# Prevalence of epilepsy in children with structural heart disease: A systematic review and meta-analysis

**DOI:** 10.64898/2026.06.27.26356733

**Authors:** Ebenezer Olatunji Adeyemi, Inimfon Akanimo Ajibola, Samuel Olu Ajigbotosho, Ayotunde Emmanuel Ajibola, Ayomide Gabriel Oladele, Julia Chigozie Okolugbo, Oluwakemi Blessing Ojulowo

**Affiliations:** Department of Paediatrics, Federal Teaching Hospital, Ido-Ekiti, Ekiti State, Nigeria; Department of Paediatrics and Child Health, Afe Babalola University, Ado-Ekiti, Nigeria; Department of Paediatrics and Child Health, Ekiti State University Teaching Hospital, Ado Ekiti, Nigeria; Emergency Department, Royal Manchester Children’s Hospital, United Kingdom

**Author notes:** **Corresponding Author** Ebenezer Olatunji Adeyemi, Department of Paediatrics, Federal Teaching Hospital, Ido-Ekiti, Ekiti State, Nigeria.

**Keywords:** congenital heart disease, structural heart disease, paediatric neurology, systematic review, meta-analysis

## Abstract

**Background:** Children with structural heart disease (SHD), particularly congenital heart disease (CHD), are increasingly recognised as being at risk of adverse neurological outcomes. Although advances in cardiac surgery and perioperative care have markedly improved survival, epilepsy has emerged as an important long-term complication. Reported prevalence estimates vary considerably across studies, and the overall burden remains uncertain. This systematic review and meta-analysis aimed to estimate the pooled prevalence of epilepsy among children with SHD and explore differences according to geographic region, lesion characteristics, and surgical exposure.

**Methods:** This systematic review and meta-analysis was conducted in accordance with PRISMA 2020 and MOOSE guidelines and registered in PROSPERO (CRD420261378572). PubMed/MEDLINE, Scopus, and ProQuest were searched for observational studies published between January 2000 and December 2025. Eligible studies included children aged 0–18 years with SHD or CHD reporting epilepsy prevalence or incidence. Two reviewers independently screened studies, extracted data, and assessed methodological quality using the Joanna Briggs Institute Critical Appraisal Checklist for Prevalence Studies. A random-effects meta-analysis was performed to estimate pooled prevalence with 95% confidence intervals (CI).

**Results:** Eight cohort studies comprising 21,731 children were included. Studies were conducted across North America, Europe, and Asia and predominantly involved surgically managed CHD populations. The pooled prevalence of epilepsy was 3.0% (95% CI 1.3%–4.8%), substantially higher than estimates reported in the general paediatric population. Heterogeneity was considerable (I² = 98.0%; p < 0.001). The 95% prediction interval ranged from 0% to 8.1%, indicating substantial variability across populations. Narrative subgroup synthesis suggested higher epilepsy prevalence among children with cyanotic and complex lesions and among surgically managed cohorts, particularly those exposed to cardiopulmonary bypass and perioperative neurological complications. Most studies were rated as having low risk of bias, and sensitivity analyses demonstrated stable findings.

**Conclusions:** Children with SHD have a substantially increased burden of epilepsy compared with the general paediatric population. Complex lesions, perioperative neurological injury, and cardiac surgical exposure may contribute to epileptogenesis. Long-term neurological surveillance and multidisciplinary neurodevelopmental follow-up should be integrated into routine care for children with SHD.

## 1. Background

Structural heart disease (SHD), particularly congenital heart disease (CHD), remains the most common congenital anomaly worldwide and a major contributor to childhood morbidity. Advances in prenatal diagnosis, surgical management, and perioperative care have substantially improved survival, such that most affected children now reach adulthood.^[1]^ Consequently, clinical focus has evolved beyond survival to encompass long-term outcomes, particularly neurodevelopmental and neurological sequelae. Among these, epilepsy has emerged as a clinically significant complication with important implications for cognitive development, quality of life, and healthcare utilisation.

Epilepsy is among the most prevalent chronic neurological disorders in childhood, with a global prevalence estimated at 5–8 per 1000 children.^[2]^ Accumulating evidence indicates that children with SHD are at an increased risk of developing epilepsy compared with the general population, with some population-based studies reporting up to a fourfold elevation in risk.^[3]^ The mechanisms underlying this association are complex and likely multifactorial. Children with SHD are exposed to a spectrum of neurological insults, including chronic hypoxia, reduced cerebral perfusion, thromboembolic phenomena, and perioperative brain injury associated with cardiopulmonary bypass and intensive care.^[4–6]^ These factors may lead to structural and functional brain abnormalities that predispose to epileptogenesis. In addition, shared genetic and developmental pathways affecting both the cardiovascular and central nervous systems may further contribute to this association.^[6]^

Despite increasing recognition of epilepsy as a comorbidity in SHD, reported prevalence estimates remain highly variable. Cohort studies have documented rates ranging from approximately 3% to over 5%, with substantial heterogeneity across populations.^[4,5]^ This variability may reflect differences in cardiac lesion type and severity, distinctions between cyanotic and acyanotic defects, variations in treatment approaches, and methodological differences between studies. Such inconsistencies limit accurate estimation of disease burden and impede the identification of high-risk subgroups.

Conceptually, the association between SHD and epilepsy can be understood within a neurocardiac interaction framework that integrates developmental and pathophysiological mechanisms linking cardiac dysfunction to neurological injury. This perspective aligns with the Multifactorial Disease Theory, which posits that disease outcomes arise from the interplay of biological, genetic, and environmental determinants rather than a single causal pathway, as shown in Figure 1.^[7]^ Within this framework, epilepsy in children with SHD is considered the result of cumulative neurological vulnerability related to underlying cardiac pathology, treatment-related exposures, and broader contextual factors.

**Figure 1.**
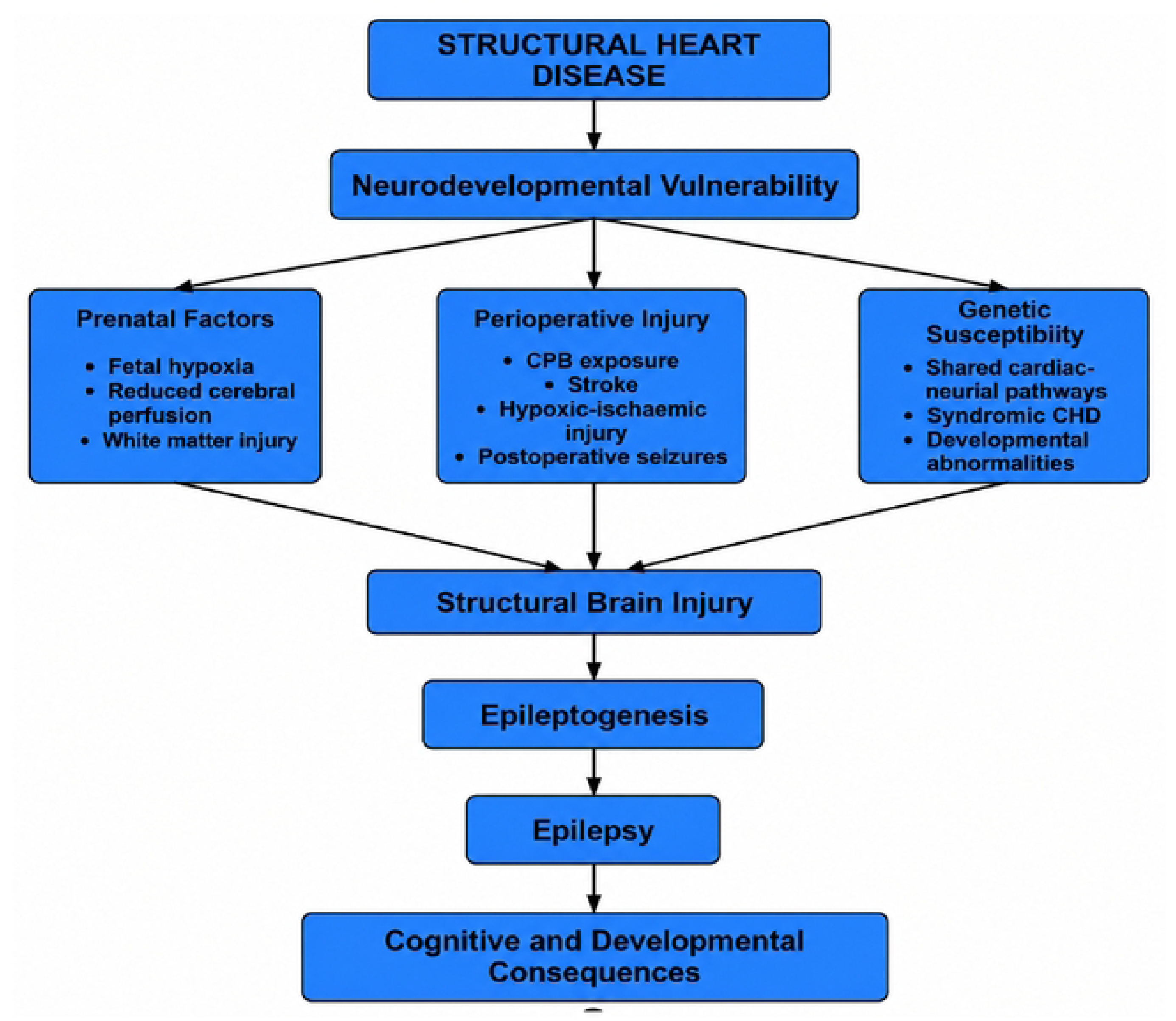
Proposed Pathways Linking Structural Heart Disease to Epilepsy and Neurodevelopmental Outcomes.

Although several cohort studies have reported an increased risk of epilepsy among children with SHD, no previous systematic review has quantitatively synthesized prevalence estimates across paediatric populations; therefore, given these uncertainties, the present study aimed to estimate the pooled prevalence of epilepsy among children with SHD through a systematic review and meta-analysis, while secondarily exploring regional differences, comparing cyanotic and acyanotic lesions, assessing the impact of surgical versus non-surgical management, investigating sources of heterogeneity, and appraising the methodological quality of included studies in accordance with PRISMA and MOOSE recommendations using the PEO framework to ensure methodological rigor and transparency.^[8–10]^

### Study Objectives

This systematic review and meta-analysis aimed to estimate the pooled prevalence of epilepsy among children with structural heart disease, including congenital heart disease. Secondary objectives were to:

1. Compare prevalence according to cyanotic versus acyanotic lesions;
2. Explore potential differences between surgically and non-surgically managed patients;
3. Explore sources of heterogeneity across studies; and
4. Assess the methodological quality of included studies

## 2. Methods

### 2.1 Study Design and Reporting Standards

This systematic review and meta-analysis were conducted in accordance with the Preferred PRISMA 2020 statement and reported following the MOOSE guidelines for observational studies. The methodology is structured using the Population–Exposure–Outcome (PEO) framework to ensure clarity and reproducibility. The protocol for the review was registered with the International Prospective Register of Systematic Reviews (PROSPERO) under registration number CRD420261378572 and published publicly on April 29, 2026.

### 2.2 Eligibility Criteria

#### 2.2.1 Inclusion Criteria

Studies were eligible if they involved paediatric populations, defined as individuals aged 0–18 years, with a confirmed diagnosis of structural heart disease (SHD), including congenital heart disease (CHD). For the purposes of this review, SHD primarily referred to congenital structural cardiac abnormalities, encompassing both simple and complex anatomical defects identified at birth or later in life. Diagnoses were required to be established through standard clinical evaluation and supported by objective evidence, such as cardiac imaging (e.g., echocardiography) and/or intraoperative findings, to ensure diagnostic consistency and reliability across included studies.

The exposure of interest is the presence of structural or anatomical heart disease as outlined above. Studies were included if SHD was clearly defined and diagnosed using recognized clinical, imaging, or surgical criteria. This inclusive approach permits representation of a wide range of cardiac lesions while preserving methodological robustness in exposure classification. The primary outcome of interest is epilepsy. Eligible studies must report either the prevalence or incidence of epilepsy within the study population. Epilepsy will be defined in accordance with established diagnostic standards, including a clinical diagnosis made by a qualified healthcare professional, classification based on the International Classification of Diseases (ICD), or the application of criteria developed by the International League Against Epilepsy (ILAE). Specifically, the definition will align with contemporary ILAE criteria, which describe epilepsy as a disorder of the brain characterised by at least two unprovoked seizures occurring more than 24 hours apart, one unprovoked seizure with a high probability of recurrence, or the diagnosis of an epilepsy syndrome.^[11,12]^

Data extraction was performed using a data collection form developed specifically for this review to ensure completeness, consistency, and reproducibility. Prior to formal data extraction, the form was piloted on a sample of eligible studies and refined as necessary to ensure clarity, consistency, and capture of all relevant variables. Two authors (EOA and IAA) independently extracted data from all included studies,

To facilitate accurate quantitative synthesis, only studies providing sufficient data to derive prevalence estimates, specifically, the number of epilepsy cases and the total sample size, were included. Eligible studies were limited to observational designs, including cross-sectional, cohort, and case–control studies, provided that sufficient data were available to extract or calculate prevalence estimates. This approach ensured the inclusion of study designs most appropriate for estimating disease frequency while maintaining methodological consistency across included studies. With respect to publication characteristics, only peer-reviewed original research articles conducted in human populations were considered. This restriction was intended to ensure the inclusion of studies with adequate methodological rigor and reliability for quantitative synthesis.

#### 2.2.2 Exclusion Criteria

Studies were excluded if they involved exclusively adult populations (>18 years) without separately reported paediatric data. Case reports and small case series comprising fewer than 10 participants were also excluded to minimise the influence of small-sample bias. Non-original publications, including reviews, editorials, commentaries, and letters lacking primary data, were not considered eligible. Conference abstracts without accessible full-text articles were excluded due to insufficient methodological detail.

Animal studies were excluded to maintain clinical relevance to human populations. Studies that did not report epilepsy as a distinct outcome, separate from other neurological conditions, were also excluded to ensure outcome specificity. Furthermore, studies focusing solely on acquired (non-structural) heart disease were not eligible for inclusion. In instances of duplicate or overlapping publications, the most comprehensive and methodologically robust dataset was retained to avoid double-counting and ensure data integrity.

### 2.3 Information Sources and Search Strategy

A comprehensive literature search was conducted across three electronic databases, including PubMed/MEDLINE, ProQuest, and Scopus, to identify relevant studies. Grey literature sources, including dissertations, conference proceedings, and unpublished registry data, were not systematically searched. There were no initial restrictions on publication year; the final search was limited to studies published between 1st January 2000 and 31st December 2025. There was no language restriction during the search. The search strategy was developed and executed by three authors (EOA, IAA, and SOA) using a combination of controlled vocabulary (e.g., MeSH terms) and free-text keywords related to “epilepsy” or “seizures,” “structural heart disease,” “congenital heart disease,” or “cardiac malformations,” and “children” or “paediatric.” Boolean operators (AND/OR) and truncation were applied to optimise the sensitivity and specificity of the search. The search was conducted on multiple dates in April 2026 (see Additional File 1 for the full search strategy). To ensure completeness, the reference lists of all included studies and relevant review articles were manually screened for additional eligible studies. Although a medical librarian specialist was not formally involved in developing the search strategy, the search syntax was iteratively refined by the review team using database-specific controlled vocabulary and free-text terms

### 2.4 Study Selection

All retrieved records were imported into the Rayyan platform, and duplicates were identified and removed before screening.^[13]^ Before formal screening, reviewers conducted a pilot calibration exercise on a subset of studies to ensure consistent interpretation of eligibility criteria. Two authors (EOA and SOA) independently screened titles, abstracts, and full-text articles for eligibility. Any disagreements were resolved through discussion and consensus and, when necessary, through consultation with a third author (IAA). Inter-reviewer agreement was substantial (Cohen’s κ = 0.76).^[14]^ The overall study selection process was documented using a PRISMA flow diagram to ensure transparency and reproducibility.

### 2.5 Data Extraction

Data extraction was performed using a data collection form developed specifically for this review to ensure completeness, consistency, and reproducibility. Prior to formal data extraction, the form was piloted on a sample of eligible studies and refined as necessary to ensure clarity, consistency, and capture of all relevant variables. Two authors (EOA and IAA) independently extracted data from all included studies, and any discrepancies were resolved through discussion and consensus, with involvement of a third author where necessary. The data extraction framework was designed to align with the study objectives and planned subgroup and meta-analytic analyses. Extracted variables included study characteristics, such as first author, year of publication, country and geographic region, study design, study setting, and, where available, funding sources and additional notes relevant to the study context.

Detailed population characteristics were recorded, including total sample size, age distribution (mean or median and range), sex distribution, and reported inclusion and exclusion criteria. For exposure assessment, data were collected on the type of SHD, including specific diagnoses, classification as cyanotic or acyanotic lesions, severity when reported, and the method used to confirm the diagnosis (e.g., echocardiography, clinical evaluation, or surgical findings). Information on associated comorbidities and genetic syndromes was also extracted when available.

For outcome measures, the number of epilepsy cases and the total study population were extracted to calculate prevalence estimates. Additional outcome-related variables included reported prevalence, age at epilepsy diagnosis, seizure type, where reported, and duration of follow-up. Relevant clinical and management variables were also extracted, including whether participants underwent surgical or non-surgical management, type and timing of surgical interventions, and subgroup-specific prevalence estimates (e.g., surgical versus non-surgical groups and cyanotic versus acyanotic lesions), where available.

In addition, key quantitative data required for meta-analysis were extracted or derived, including the number of epilepsy cases, total sample size, calculated proportions, and corresponding measures of uncertainty (e.g., standard errors and confidence intervals). Risk-of-bias–related variables were also recorded to support quality assessment. This comprehensive and structured approach ensured that all relevant data were captured to facilitate robust synthesis, subgroup analyses, and exploration of heterogeneity.

### 2.6 Risk of Bias Assessment

Methodological quality and risk of bias of the included studies were appraised independently by two authors (EOA and IAA) using the Joanna Briggs Institute (JBI) Critical Appraisal Checklist for Studies Reporting Prevalence Data.^[15]^ Each item on the 9-question checklist was scored dichotomously, where a’Yes’ (Y) response was awarded 1 point, and a’No’ (N),’Unclear’ (U), or’Not Applicable’ (N/A) response was awarded 0 points. Based on the total cumulative score, the risk of bias for each study was stratified as Low Risk of Bias (≥7’Yes’ answers), Moderate Risk of Bias (5 - 6 ‘Yes’ answers), or High Risk of Bias (≤ 4’Yes’ answers). Although several included primary studies utilized longitudinal or cohort designs, the JBI prevalence tool was selected as the primary assessment instrument because the overarching objective of this systematic review is to synthesize and estimate an absolute pooled prevalence of epilepsy, rather than to establish relative risk ratios or causal associations. Where primary studies featured longitudinal cohorts, criteria regarding sampling frame representativeness and recruitment were mapped to the baseline cohort selection, and response rate assessments (Question 9 on the JBI) were evaluated based on participant attrition and loss to follow-up over the study period. Discrepancies in scoring were resolved via consensus or consultation with a third author.

### 3.7 Data Synthesis and Statistical Analysis

#### 3.7.1 Pooled Prevalence Estimation

Studies rated as having low or moderate risk of bias were included in the meta-analysis and used for the overall synthesis of findings. The data were analyzed using Review Manager 5.4.1 (The Nordic Cochrane Centre, Copenhagen, Denmark). A random-effects meta-analysis was performed to estimate the pooled prevalence of epilepsy among children with structural heart disease. Prevalence estimates from individual studies were converted to proportions and pooled with their corresponding 95% confidence intervals. A random-effects model was selected because substantial clinical and methodological heterogeneity between studies was anticipated.

#### 3.7.2 Heterogeneity Assessment

Statistical heterogeneity across studies was assessed using Cochran’s Q test and quantified using the I² statistic.^[16]^ I² values of 25%, 50%, and 75% were interpreted as low, moderate, and high heterogeneity, respectively. A statistically significant Q statistic (p < 0.10) was considered indicative of significant heterogeneity.

#### 3.7.3 Subgroup Analyses

Prespecified subgroup analyses were conducted to explore potential sources of heterogeneity. Subgroups included:

- geographic region (North America, Europe, Asia),
- cyanotic versus acyanotic congenital heart disease,
- complex versus simple lesions, and
- surgically versus non-surgically managed patients.

#### 3.7.4 Meta-regression

Meta-regression was intentionally not performed, in accordance with the methodological guidelines outlined in the Cochrane Handbook for Systematic Reviews, which recommend a minimum baseline of 10 primary studies per covariate to prevent model overfitting and maintain adequate statistical power.^[17]^

#### 3.7.5 Sensitivity Analyses

Sensitivity analyses were performed to assess the robustness and stability of the pooled prevalence estimates. A leave-one-out approach was used, with each study sequentially removed from the analysis to determine its influence on the overall findings. Also, sensitivity analyses were conducted on studies rated as having a moderate risk of bias.

#### 3.7.6 Publication Bias

Publication bias was assessed visually using funnel plots.^[18]^ Due to the relatively small number of included studies, formal statistical tests for funnel plot asymmetry were interpreted cautiously.

### 3.8 Software

All statistical analyses were performed using Review Manager (RevMan) version 5.4.1 and R statistical software (meta and metafor packages). Forest plots and subgroup analyses were generated using random-effects models with corresponding 95% confidence intervals.

### 3.9 Certainty of Evidence

The certainty of evidence for the pooled prevalence estimate was independently assessed by 2 authors (EOA and IAA) using the GRADE framework adapted for prevalence studies, considering study limitations, consistency of findings, precision of estimates, indirectness, and publication bias.^[19]^

## 3. Results

### 3.1 Study Selection

A total of 642 records were identified through database searching and manual reference screening. After removal of 72 duplicates, titles and abstracts were screened for eligibility. Full-text review was subsequently conducted for potentially eligible studies. Eight studies met the eligibility criteria and were included in the systematic review and meta-analysis. The study selection process is summarized in the PRISMA flow diagram (Figure 2).

**Figure 2.**
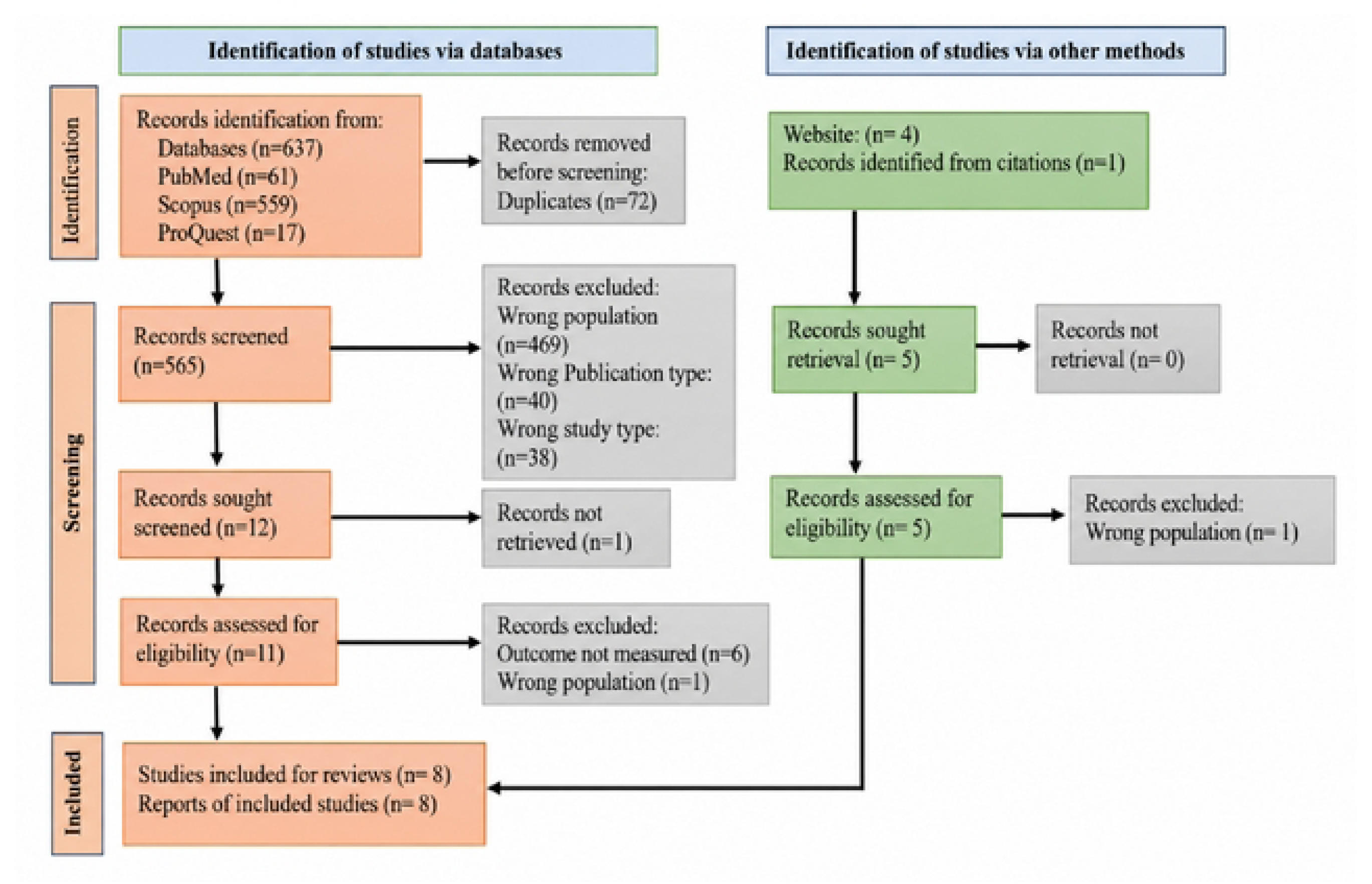
PRISMA flow diagram of study selection process

**Figure 3.**
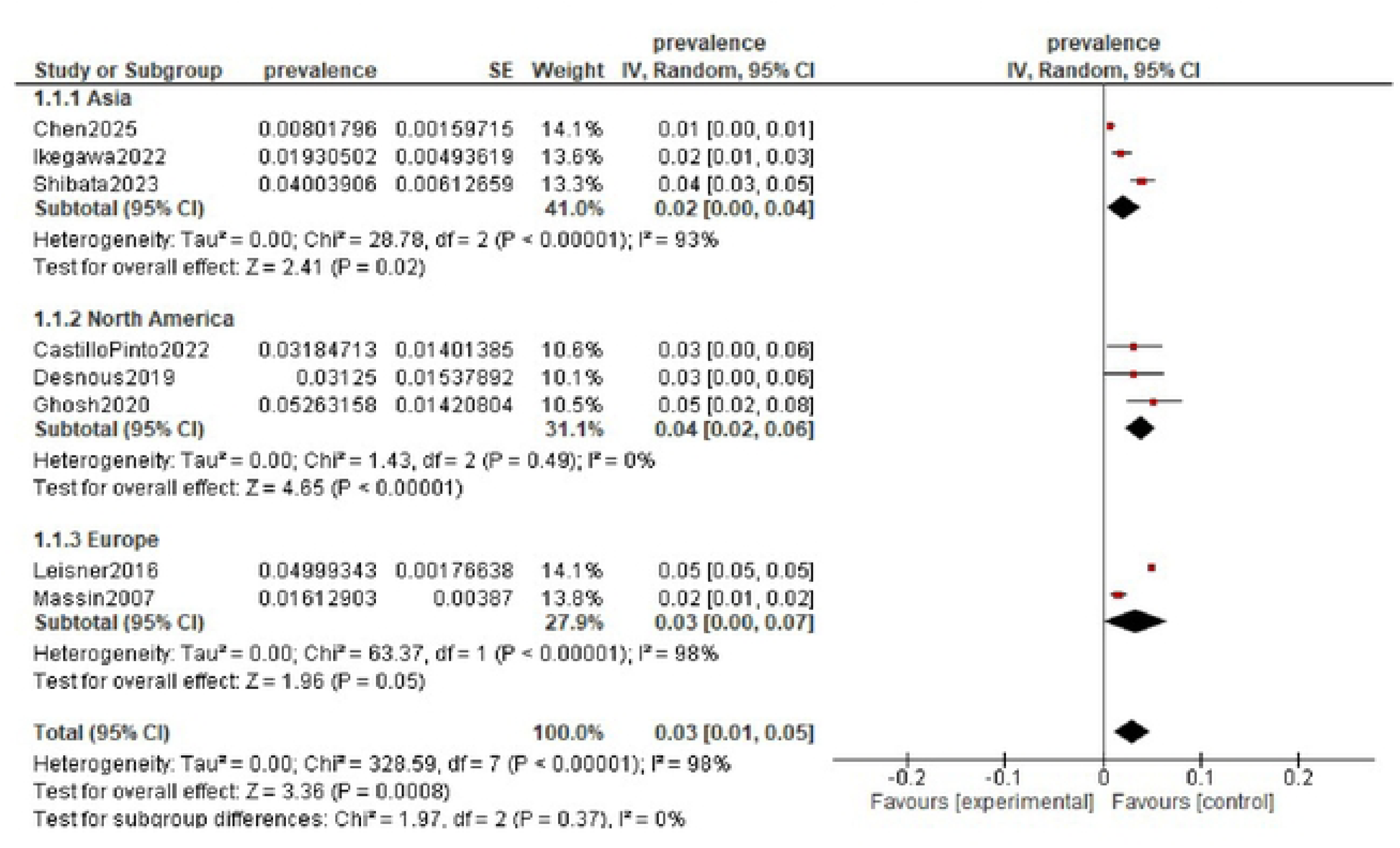
Forest plot showing the pooled and subgroup prevalences of epilepsy among children with structural heart disease from Asia, North America, and Europe.

**Figure 4.**
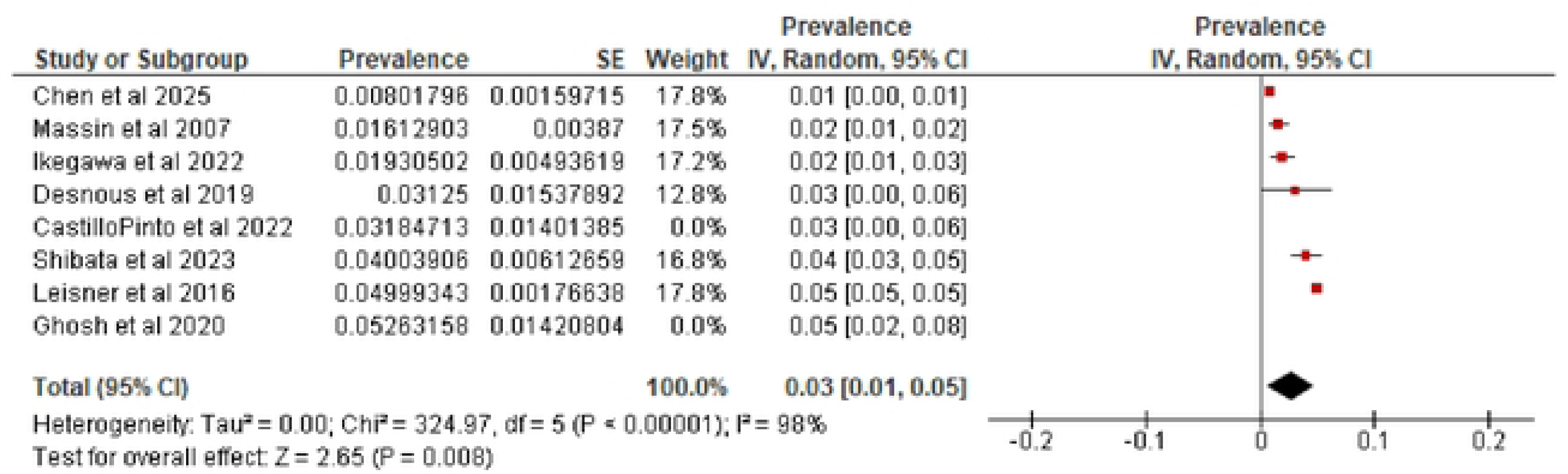
Leave-one-out sensitivity analysis

The PRISMA flow diagram illustrates the identification, screening, eligibility assessment, and final inclusion of studies in accordance with PRISMA 2020 reporting guidelines.^[8]^

### 3.2 Characteristics of Included Studies

The eight included studies were published between 2007 and 2025 and collectively enrolled 21,731 children with structural or congenital heart disease. Included studies were conducted across three geographic regions: North America (United States^[4,5]^ and Canada^[20]^), Europe (Belgium^[21]^ and Denmark^[22]^), and Asia (China^[23]^ and Japan^[6,24]^). Study designs predominantly consisted of cohort studies, including prospective cohorts,^[20,22]^ retrospective cohorts,^[6,21,23,24]^ and population-based registry cohorts^[20,22]^.

Sample sizes varied substantially across studies, ranging from 128 participants in the Canadian neurocardiac cohort reported by Desnous et al.^[20]^ to 15,222 participants in the nationwide Danish registry cohort described by Leisner et al.^[22]^ Most studies focused on children undergoing cardiac surgery, particularly those requiring cardiopulmonary bypass, while others included broader CHD populations identified through national registries or cardiac centres.

The prevalence of epilepsy across studies ranged from 0.8%^[23]^ to 5.2%^[4]^. Nearly all the studies evaluated epilepsy using clinical diagnoses, International Classification of Diseases (ICD) coding systems, or specialist neurological assessment. Several studies also explored risk factors associated with epilepsy development, including perioperative seizures, stroke, complex cyanotic lesions, prolonged intensive care admission, and cardiopulmonary bypass exposure.

**Table 1:**
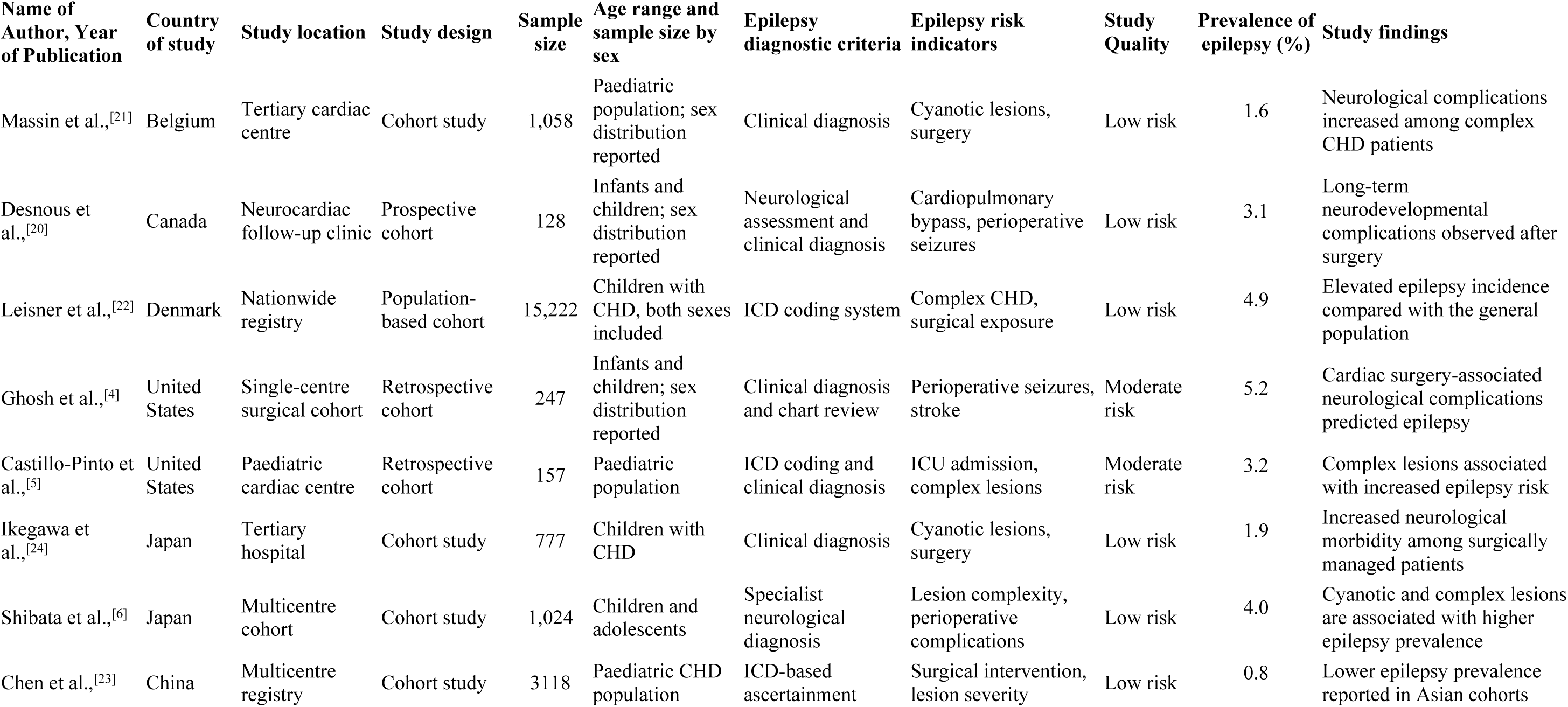
Characteristics and Some Findings of the Included Studies.

### 3.2 Risk of Bias Assessment

**Table 2.**
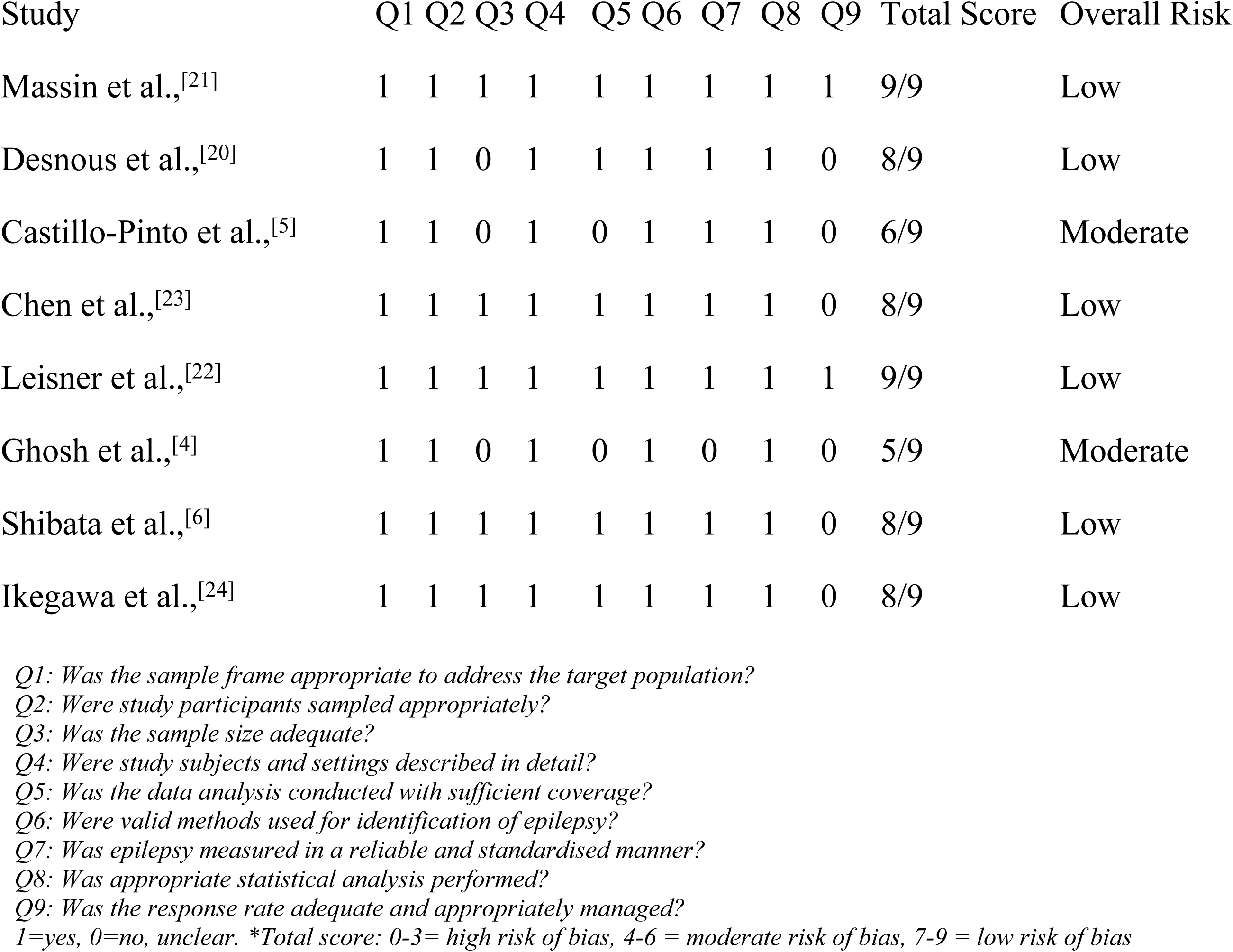
Risk of Bias Assessment of Included Studies Using the Joanna Briggs Institute (JBI) Critical Appraisal Checklist for Studies Reporting Prevalence Data.

Methodological quality assessment using the JBI Critical Appraisal Checklist for Studies Reporting Prevalence Data demonstrated generally good methodological quality across included studies. Six studies were rated as having low risk of bias, while two studies were classified as having moderate risk of bias. No study was categorized as high risk of bias.

Most studies demonstrated appropriate sampling frames, valid methods for identifying epilepsy, reliable outcome measurement, and appropriate statistical analyses. The primary methodological limitations involved incomplete sample coverage and concerns regarding participant attrition or incomplete longitudinal follow-up in retrospective hospital-based cohorts, factors known to increase the risk of selection and attrition bias in observational prevalence studies.^[15,19,25]^

Given the longitudinal design of the included cohorts, item 9 (response rate) was operationalized as the patient retention rate and the strategic management of attrition over time. These studies (Massin et al.,^[21]^; Desnous et al.,^[20]^; Chen et al.,^[1]^; Leisner et al.,^[22]^; Shibata et al., ^[6]^; and Ikegawa et al.,^[24]^) achieved a low risk of bias by maintaining complete registry tracking or documenting high retention rates alongside appropriate statistical censoring. Conversely, two retrospective cohorts (Castillo-Pinto et al.,^[5]^; Ghosh et al.,^[4]^) were scored as “No” due to high or poorly defined attrition rates resulting from missing longitudinal chart data.

### 3.3 Pooled Prevalence of Epilepsy

A random-effects meta-analysis of the eight included studies demonstrated a pooled prevalence of epilepsy of 3.0% (95% CI 1.0%–5.0%) among children with structural heart disease.

Considerable heterogeneity was observed across studies (Cochran Q P < 0.001; I² = 98.0%), supporting the use of a random-effects model. This variability likely reflected differences in study design, patient characteristics, lesion complexity, duration of follow-up, and methods used to identify epilepsy.

To facilitate interpretation of this substantial heterogeneity, a 95% prediction interval was calculated. The prediction interval ranged from approximately 0% to 8.1%, indicating that the prevalence of epilepsy in future comparable populations of children with structural heart disease may reasonably be expected to fall within this range. The broad prediction interval suggests that the observed heterogeneity reflects genuine between-population differences rather than random variation alone.

The largest prevalence estimate was reported by Ghosh et al.^[4]^ among infants undergoing cardiopulmonary bypass surgery, whereas the lowest prevalence was observed in the Chinese multicentre cohort by Chen et al.^[23]^

### 3.4 Subgroup Analyses

Prespecified subgroup analyses were conducted to evaluate differences in epilepsy prevalence according to geographic region, lesion characteristics, and treatment approach.

### Geographic Region

Subgroup analysis by geographic region demonstrated variability in epilepsy prevalence across continents.

Studies from Asia demonstrated the lowest pooled prevalence estimate (∼ 2%). However, important variability remained between studies depending on lesion complexity and surgical exposure. North America demonstrated the highest pooled prevalence estimate, approximately 4.0%, largely driven by cohorts involving children with complex congenital heart disease undergoing surgery and prolonged postoperative follow-up, while European studies demonstrated a pooled prevalence of approximately 3.0%. The largest Danish nationwide registry cohort contributed substantially to this group and provided long-term population-based follow-up data.

### Cyanotic vs Acyanotic Congenital Heart Disease

Studies that compared cyanotic and acyanotic lesions generally found a higher prevalence of epilepsy among children with cyanotic congenital heart disease.^[6,20,22]^ Formal pooled subgroup meta-analysis was limited by inconsistent subgroup-specific reporting across studies; therefore, subgroup findings were synthesized narratively (see Table 3).

**Table 3:** Findings From Prespecified Subgroup Analyses.

| Subgroup Category | Findings | Studies Reporting Findings |
| --- | --- | --- |
| Cyanotic congenital heart disease | Higher prevalence of epilepsy was consistently reported among children with cyanotic lesions. | Massin et al.,<sup>[21]</sup>; Ikegawa et al.,<sup>[24]</sup>; Shibata et al.,<sup>[6]</sup> |
| Acyanotic congenital heart disease | Lower prevalence estimates were generally observed compared with cyanotic lesions. | Massin et al.,<sup>[21]</sup>; Shibata et al.,<sup>[6]</sup> |
| Complex structural heart disease | Children with complex lesions demonstrated higher epilepsy prevalence across most studies. | Leisner et al.,<sup>[22]</sup>; Castillo-Pinto et al.,<sup>[5]</sup>; Shibata et al.,<sup>[6]</sup>; Chen et al.,<sup>[23]</sup> |
| Simple structural heart disease | Lower epilepsy prevalence was observed among children with simple lesions. | Leisner et al.,<sup>[22]</sup>; Chen et al.,<sup>[23]</sup> |
| Surgically managed patients | Surgically treated cohorts demonstrated higher epilepsy prevalence, particularly after neonatal or infant cardiac surgery. | Desnous et al.,<sup>[20]</sup>; Ghosh et al.,<sup>[4]</sup>; Ikegawa et al.,<sup>[24]</sup>; Shibata et al.,<sup>[6]</sup> |
| Non-surgically managed patients | Lower prevalence estimates were generally reported in non-surgically managed cohorts. | Leisner et al.,<sup>[22]</sup>; Chen et al.,<sup>[23]</sup> |

### Complex vs Simple Structural Heart Disease

Children with complex structural heart disease consistently appeared to have a greater burden of epilepsy than those with simpler lesions.^[5,6,22,23]^ Across both registry-based and hospital-based cohorts, more severe cardiac disease was associated with increased long-term neurological morbidity. Because subgroup-specific prevalence estimates were inconsistently reported, findings for lesion complexity were synthesized narratively rather than through formal pooled subgroup meta-analysis.

### Surgically vs Non-surgically Managed Patients

Higher epilepsy prevalence was generally observed among surgically managed patients compared with children managed without surgery. The greatest burden was reported in cohorts undergoing neonatal or early infant cardiac surgery, particularly procedures involving cardiopulmonary bypass. Perioperative seizures, stroke, and prolonged intensive care admissions were frequently identified as possible contributors to later epilepsy development. Formal pooled subgroup analysis was not feasible because detailed subgroup-level prevalence data were unavailable in several studies.

### 3.5 Sensitivity Analyses

The pooled prevalence of epilepsy remained largely unchanged at approximately 3% throughout these analyses, indicating that no individual study disproportionately influenced the primary results. The only notable change occurred after exclusion of the large population-based registry study by Leisner et al. ^[22]^, which resulted in a modest reduction in the pooled prevalence estimate to approximately 2%.

Additional sensitivity analyses were conducted by excluding studies rated as having a moderate risk of bias. Removal of the retrospective cohorts by Ghosh et al.^[4]^ and Castillo-Pinto et al.^[5]^ produced minimal change in the pooled prevalence estimate, which remained stable at approximately 3% (Figure 6). These findings suggest that the overall results were robust and were not substantially influenced by studies with greater methodological limitations.

### 3.6 Publication Bias

Visual inspection of the funnel plot suggested mild asymmetry, although interpretation was limited by the relatively small number of included studies. Consequently, formal statistical testing for publication bias was interpreted cautiously.

### 3.7 Interpretation of Heterogeneity

The substantial heterogeneity observed across included studies was anticipated given the marked clinical diversity of congenital and structural heart disease populations. Included cohorts differed in lesion complexity, proportions of cyanotic defects, surgical exposure, duration of follow-up, epilepsy ascertainment methods, and healthcare settings. Such variability is common in prevalence meta-analyses and may reflect true differences in disease burden rather than methodological inconsistency alone.

Importantly, despite the high I² statistic, all studies demonstrated a prevalence of epilepsy exceeding that typically reported in the general paediatric population. Furthermore, sensitivity analyses showed stable pooled estimates following exclusion of individual studies and studies at moderate risk of bias. Together, these findings suggest that while the exact magnitude of epilepsy burden may vary across populations, the overall conclusion that children with structural heart disease are at increased risk of epilepsy remains robust

### 3.8 Certainty of Evidence

Using GRADE principles adapted for prevalence studies, the overall certainty of evidence was judged to be moderate.^[19]^

**Table 4.**
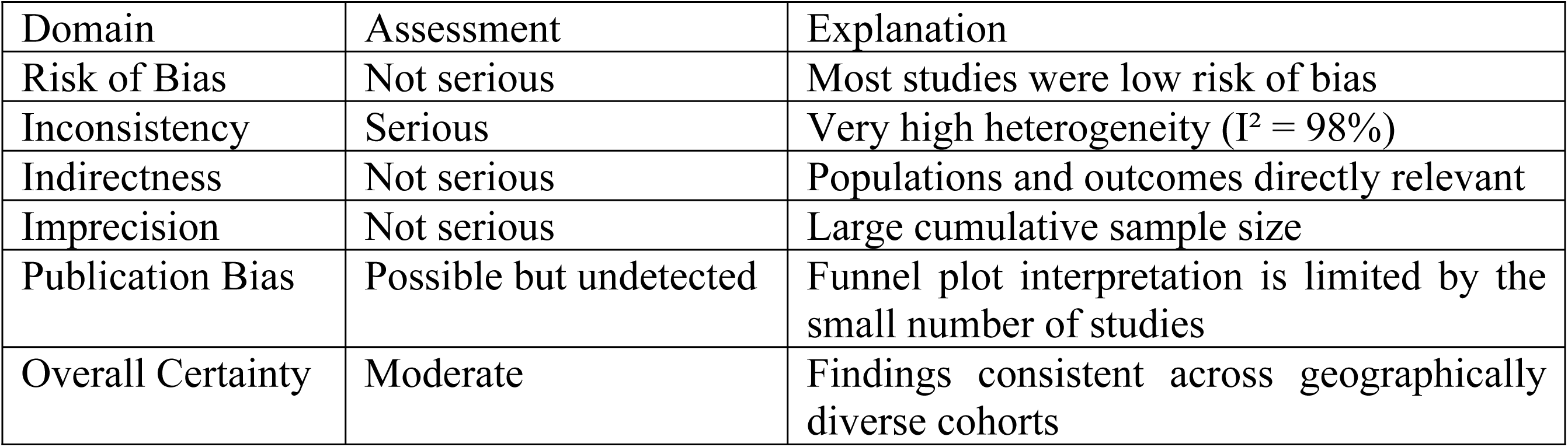
GRADE Evidence Profile.

| Domain | Assessment | Explanation |
| --- | --- | --- |
| Risk of Bias | Not serious | Most studies were low risk of bias |
| Inconsistency | Serious | Very high heterogeneity ($I^2 = 98\%$) |
| Indirectness | Not serious | Populations and outcomes directly relevant |
| Imprecision | Not serious | Large cumulative sample size |
| Publication Bias | Possible but undetected | Funnel plot interpretation is limited by the small number of studies |
| Overall Certainty | Moderate | Findings consistent across geographically diverse cohorts |

Although inconsistency was substantial, confidence in the overall estimate was strengthened by the large cumulative sample size, predominance of low-risk studies, stability of sensitivity analyses, and consistency of the direction of association across all included cohorts.

## 4. Discussion

This systematic review and meta-analysis found that epilepsy is an important neurological comorbidity among children with SHD. While the prevalence of epilepsy in children is estimated at approximately 5–8 per 1,000 population (0.5–0.8%), among children with SHD, the prevalence is about 3 per 100 people, representing a substantially greater neurological burden.^[2]^ These findings are consistent with previous evidence demonstrating increased neurological and neurodevelopmental vulnerability among children with congenital heart disease (CHD).^[3,26,27]^ In the present review, North American studies demonstrated higher pooled prevalence estimates than studies from Asia, although substantial heterogeneity remained within all geographic regions.

The relationship between SHD and epilepsy is increasingly recognised as part of a broader neurodevelopmental phenotype rather than an isolated neurological complication. Contemporary neuroimaging and developmental studies have demonstrated that abnormalities of brain maturation may be present before birth in fetuses with congenital heart disease. Reduced cerebral oxygen delivery, altered fetal cerebral blood flow, delayed cortical maturation, and impaired white matter development have all been described in this population and may establish a substrate of neurological vulnerability that persists throughout childhood.^[26,27]^ These prenatal abnormalities may partially explain why neurological and developmental impairments are frequently observed even before exposure to cardiac surgery or other major postnatal interventions

Following birth, additional physiological and treatment-related insults may further increase susceptibility to epileptogenesis. Across the included studies, chronic hypoxaemia, impaired cerebral perfusion, thromboembolic complications, haemodynamic instability, perioperative neurological injury, and shared neurodevelopmental vulnerabilities were identified as potential contributors to epilepsy development.^[4–6]^ Children with complex cyanotic lesions appeared particularly vulnerable, likely because of repeated exposure to physiological instability, cardiopulmonary bypass, and invasive interventions during critical periods of brain development. Cardiac surgery involving cardiopulmonary bypass may expose the developing brain to inflammatory activation, microembolic phenomena, reperfusion injury, and fluctuations in cerebral blood flow, all of which may contribute to the formation of epileptogenic networks and long-term seizure disorders.

The findings of the present review support this mechanistic framework. Higher epilepsy prevalence was consistently observed among children with complex lesions, cyanotic defects, and surgically managed cohorts. These populations are more likely to experience cumulative neurological insults and, therefore, may represent groups at particularly elevated risk of epilepsy. The observed association between lesion complexity and epilepsy burden suggests that epilepsy may represent an important marker of overall neurological vulnerability within the congenital heart disease population rather than merely a consequence of isolated perioperative events.

Shared developmental pathways may provide an additional explanation for the association between SHD and epilepsy. Cardiac and cerebral development occur concurrently during embryogenesis and involve overlapping genetic and molecular signalling pathways. Consequently, genetic abnormalities affecting cardiovascular morphogenesis may simultaneously influence neuronal migration, cortical organisation, synaptic development, and seizure susceptibility. This concept is supported by the increased prevalence of neurodevelopmental disorders observed among children with congenital heart disease, even in the absence of major perioperative complications. ^[26,27]^ Together, these observations support a multifactorial model of epileptogenesis in SHD, whereby prenatal brain dysmaturation, acquired neurological injury, treatment-related exposures, and genetic susceptibility interact to increase long-term epilepsy risk.

The present findings add to the growing body of literature describing adverse neurodevelopmental outcomes among survivors of CHD. Previous studies have demonstrated increased rates of cognitive impairment, developmental delay, stroke, and seizure disorders among children with complex congenital lesions.^[26,27]^ The findings of this review further underscore the importance of epilepsy as part of the broader spectrum of neurological morbidity associated with SHD and support recommendations for long-term neurological surveillance and integrated neurodevelopmental follow-up in this population.

One methodological challenge encountered during this review involved adapting the JBI Critical Appraisal Checklist for Prevalence Data to the predominantly longitudinal cohort studies included in the analysis. In particular, assessment of “response rates” (Question 9) required consideration of participant retention and the management of loss to follow-up over time. Population-based registry studies generally demonstrated excellent follow-up and minimal missing data, whereas retrospective hospital-based cohorts were more susceptible to incomplete longitudinal records, which may have introduced some bias in estimating the long-term cumulative prevalence of epilepsy in children with SHD.

Considerable heterogeneity was observed across studies (I² = 98%), likely reflecting differences in lesion severity, duration of follow-up, epilepsy ascertainment methods, perioperative neurological surveillance, geographic setting, and the proportion of surgically managed patients. Accordingly, the pooled prevalence estimate should be interpreted cautiously, as it represents an average across clinically and methodologically diverse populations rather than a single universally applicable prevalence value. Nevertheless, the calculated prediction interval suggested that epilepsy prevalence in future comparable populations would likely remain higher than estimates reported for the general paediatric population. Furthermore, sensitivity analyses demonstrated stable pooled estimates following sequential study exclusion and removal of studies at moderate risk of bias, supporting the robustness of the overall findings despite substantial between-study variability.

These findings have important clinical implications. Given that approximately one in thirty children with SHD may develop epilepsy, structured neurological surveillance should be considered an integral component of long-term congenital heart disease care. Particular attention should be directed towards children with complex lesions, cyanotic heart disease, perioperative neurological complications, or a history of postoperative seizures. Multidisciplinary follow-up programmes involving paediatric cardiologists, neurologists, developmental paediatricians, intensivists, and rehabilitation specialists may facilitate earlier identification of seizure disorders and optimise neurodevelopmental outcomes. The findings also support the incorporation of epilepsy screening into established neurodevelopmental follow-up pathways for survivors of complex congenital heart disease.

Several limitations should be acknowledged. First, substantial heterogeneity was observed across studies (I² = 98%), and therefore, the pooled prevalence estimate should be interpreted cautiously as an average across diverse populations rather than a precise estimate applicable to all children with structural heart disease, likely reflecting differences in study design, patient characteristics, lesion severity, epilepsy definitions, and duration of follow-up. Some studies relied on administrative coding systems, which may underestimate or misclassify epilepsy diagnoses.^[5,6]^ Additionally, relatively few studies provided detailed subgroup-specific data for cyanotic versus acyanotic lesions or surgical versus non-surgical cohorts, limiting quantitative subgroup analyses. Grey literature sources, including theses, conference abstracts, and unpublished registry data, were not systematically searched, potentially increasing susceptibility to publication bias. Furthermore, the literature search was restricted to PubMed/MEDLINE, Scopus, and ProQuest; although reference lists of included studies and relevant reviews were manually screened to identify additional eligible studies, the omission of databases such as Embase and Web of Science may have reduced the comprehensiveness of study identification. Resource constraints influenced database selection. Most included studies also originated from high-income countries, which may limit the generalisability of the findings to low-and middle-income settings where access to specialised cardiac and neurological care differs substantially.

Despite these limitations, this study provides the first comprehensive quantitative synthesis of epilepsy prevalence among children with structural heart disease. The large cumulative sample size, inclusion of geographically diverse cohorts, generally favourable methodological quality, and robustness of sensitivity analyses strengthen confidence in the findings. Overall, the evidence indicates that children with SHD represent a neurologically vulnerable population with a substantially elevated burden of epilepsy, highlighting the need for integrated neurodevelopmental surveillance and long-term multidisciplinary care.

### Clinical Implications

The present findings have important implications for the long-term management of children with structural heart disease. Given that approximately one in thirty affected children may develop epilepsy, routine neurodevelopmental surveillance should be considered an integral component of congenital heart disease care. Particular attention should be directed towards children with complex lesions, cyanotic heart disease, perioperative neurological complications, or a history of postoperative seizures.

Multidisciplinary follow-up programmes involving paediatric cardiologists, neurologists, developmental paediatricians, and rehabilitation specialists may facilitate earlier identification of seizure disorders and optimise neurodevelopmental outcomes. The findings also support the incorporation of epilepsy screening into established neurodevelopmental follow-up pathways for survivors of complex congenital heart disease.

### Future Research

Future studies should investigate lesion-specific epilepsy risk, identify genetic determinants of epileptogenesis in congenital heart disease, and evaluate the contribution of perioperative neurological injury to long-term seizure outcomes. Large multicentre prospective cohorts with standardised neurological assessment and neuroimaging protocols are needed to clarify causal mechanisms and identify opportunities for neuroprotective interventions.

Further research should also evaluate whether structured neurological surveillance programmes improve early epilepsy detection, treatment initiation, neurodevelopmental outcomes, and quality of life among children with structural heart disease.

## 5. Conclusion

Children with structural heart disease appear to have a substantially greater burden of epilepsy than the general paediatric population. Complex congenital lesions, perioperative neurological injury, and exposure to cardiac surgery may contribute to increased susceptibility to epileptogenesis. These findings underscore the importance of integrating long-term neurological surveillance and multidisciplinary care into routine management for children with structural heart disease

## List of abbreviations

CHD: Congenital heart disease
SHD: Structural heart disease
CI: Confidence interval
ECMO: Extracorporeal membrane oxygenation
GRADE: Grading of Recommendations Assessment, Development and Evaluation
ICD: International Classification of Diseases
ILAE: International League Against Epilepsy
ICU: Intensive care unit
JBI: Joanna Briggs Institute
MOOSE: Meta-analysis of Observational Studies in Epidemiology
PEO: Population–Exposure–Outcome
PRISMA: Preferred Reporting Items for Systematic Reviews and Meta-analyses
PROSPERO: International Prospective Register of Systematic Reviews
RevMan: Review Manager
RR: Risk ratio

## Declarations

### Ethics approval and consent to participate

Not applicable

### Consent for publication

Not applicable

### Availability of data and materials

All data used in the study are available to the public

### Competing interests

All the authors have no competing interests to declare

### Funding

Not applicable

### Authors’ contributions

EOA: Conceptualization, protocol review, searched, screened titles and abstracts, full text screening, extracted data, risk of bias assessment, conducted the meta-analysis, drafted the manuscript, and reviewed the manuscript for intellectual inputs.

IAA: Conceptualization, protocol review, generated search terms, searched, full text screening, risk of bias assessment, and reviewed the manuscript for intellectual inputs.

SOA: Protocol review, generated search terms, searched, screened titles and abstracts, calculated interrater reliability, and reviewed the manuscript for intellectual inputs.

AEA: Protocol review, and reviewed the manuscript for intellectual inputs

AGO: Protocol review, and reviewed the manuscript for intellectual inputs

JCO: Protocol review, and reviewed the manuscript for intellectual inputs

OBO: Protocol review, generated search terms, searched, meta-analysis, manuscript review for intellectual input, technical support, and project management.

## Data Availability

No new data were generated in this study. All data analyzed were extracted from previously published studies included in this systematic review and meta-analysis. All primary studies included in the review are cited in the reference list.

## Acknowledgments

The authors thank the Research Development Hub for the training platform in systematic review and meta-analysis to advance research capacity in Africa.

---

**Supplementary File 1:**
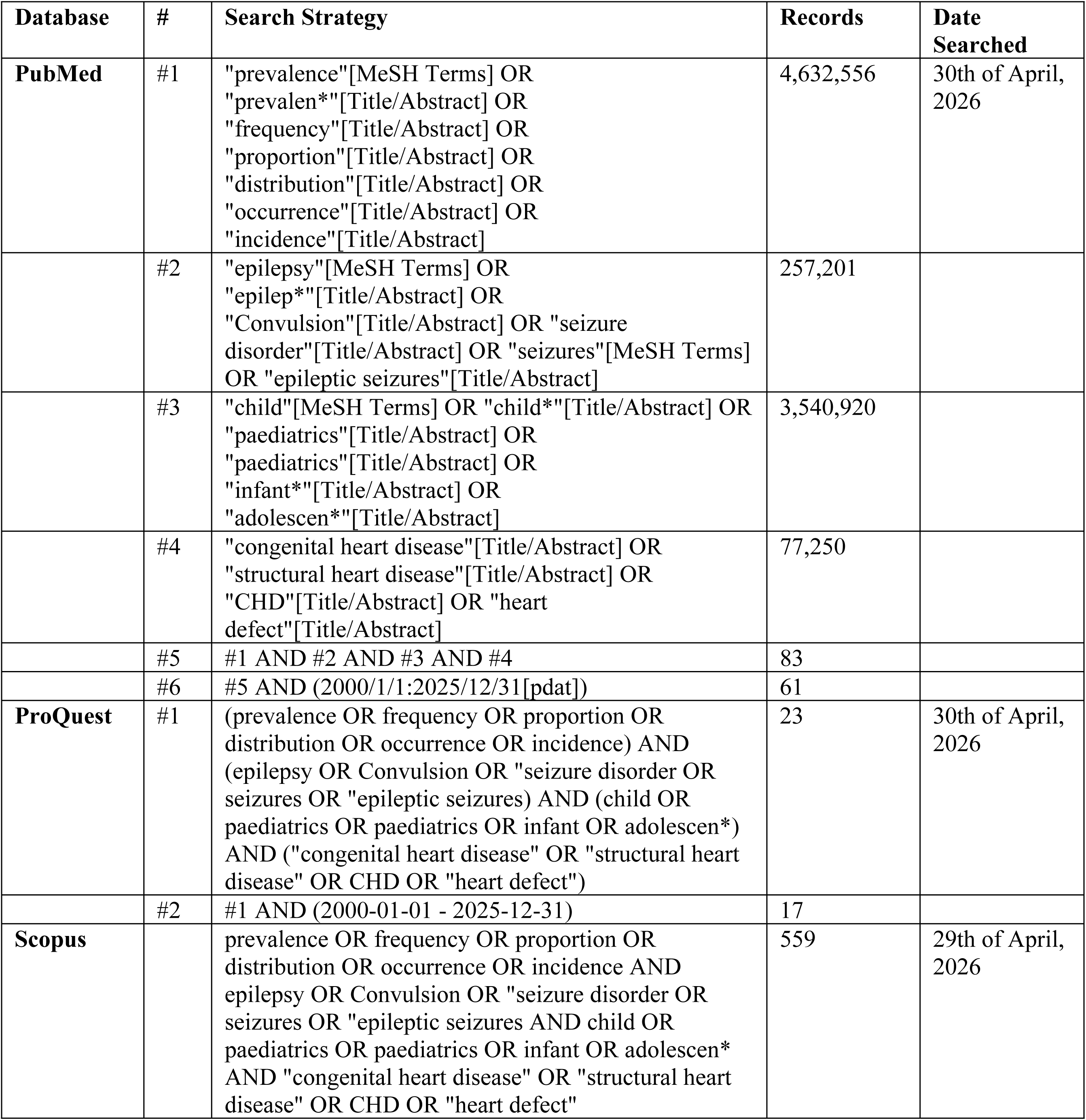
Search Strategy.

| Database | # | Search Strategy | Records | Date Searched |
| --- | --- | --- | --- | --- |
| PubMed | #1 | "prevalence"[MeSH Terms] OR<br>"prevalen*"[Title/Abstract] OR<br>"frequency"[Title/Abstract] OR<br>"proportion"[Title/Abstract] OR<br>"distribution"[Title/Abstract] OR<br>"occurrence"[Title/Abstract] OR<br>"incidence"[Title/Abstract] | 4,632,556 | 30th of April, 2026 |
|  | #2 | "epilepsy"[MeSH Terms] OR<br>"epilep*"[Title/Abstract] OR<br>"Convulsion"[Title/Abstract] OR "seizure disorder"[Title/Abstract] OR "seizures"[MeSH Terms] OR "epileptic seizures"[Title/Abstract] | 257,201 |  |
|  | #3 | "child"[MeSH Terms] OR "child*"[Title/Abstract] OR<br>"paediatrics"[Title/Abstract] OR<br>"paediatrics"[Title/Abstract] OR<br>"infant*"[Title/Abstract] OR<br>"adolescen*"[Title/Abstract] | 3,540,920 |  |
|  | #4 | "congenital heart disease"[Title/Abstract] OR<br>"structural heart disease"[Title/Abstract] OR<br>"CHD"[Title/Abstract] OR "heart defect"[Title/Abstract] | 77,250 |  |
|  | #5 | #1 AND #2 AND #3 AND #4 | 83 |  |
|  | #6 | #5 AND (2000/1/1:2025/12/31[pdat]) | 61 |  |
| ProQuest | #1 | (prevalence OR frequency OR proportion OR distribution OR occurrence OR incidence) AND (epilepsy OR Convulsion OR "seizure disorder OR seizures OR "epileptic seizures) AND (child OR paediatrics OR paediatrics OR infant OR adolescen*) AND ("congenital heart disease" OR "structural heart disease" OR CHD OR "heart defect") | 23 | 30th of April, 2026 |
|  | #2 | #1 AND (2000-01-01 - 2025-12-31) | 17 |  |
| Scopus |  | prevalence OR frequency OR proportion OR distribution OR occurrence OR incidence AND epilepsy OR Convulsion OR "seizure disorder OR seizures OR "epileptic seizures AND child OR paediatrics OR paediatrics OR infant OR adolescen* AND "congenital heart disease" OR "structural heart disease" OR CHD OR "heart defect" | 559 | 29th of April, 2026 |

**Supplementary File 2:**
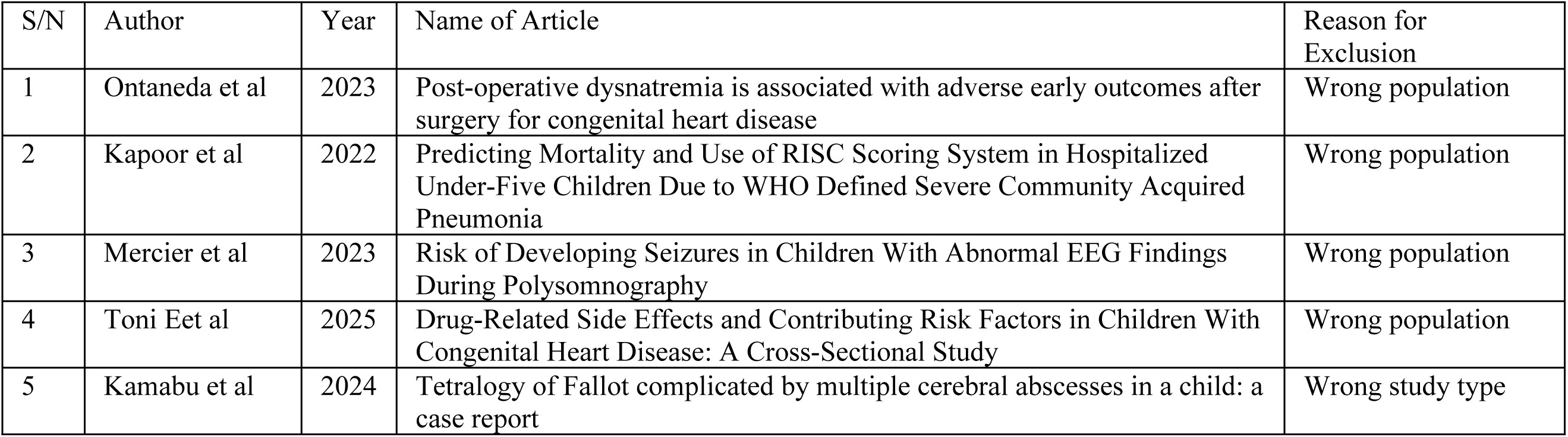
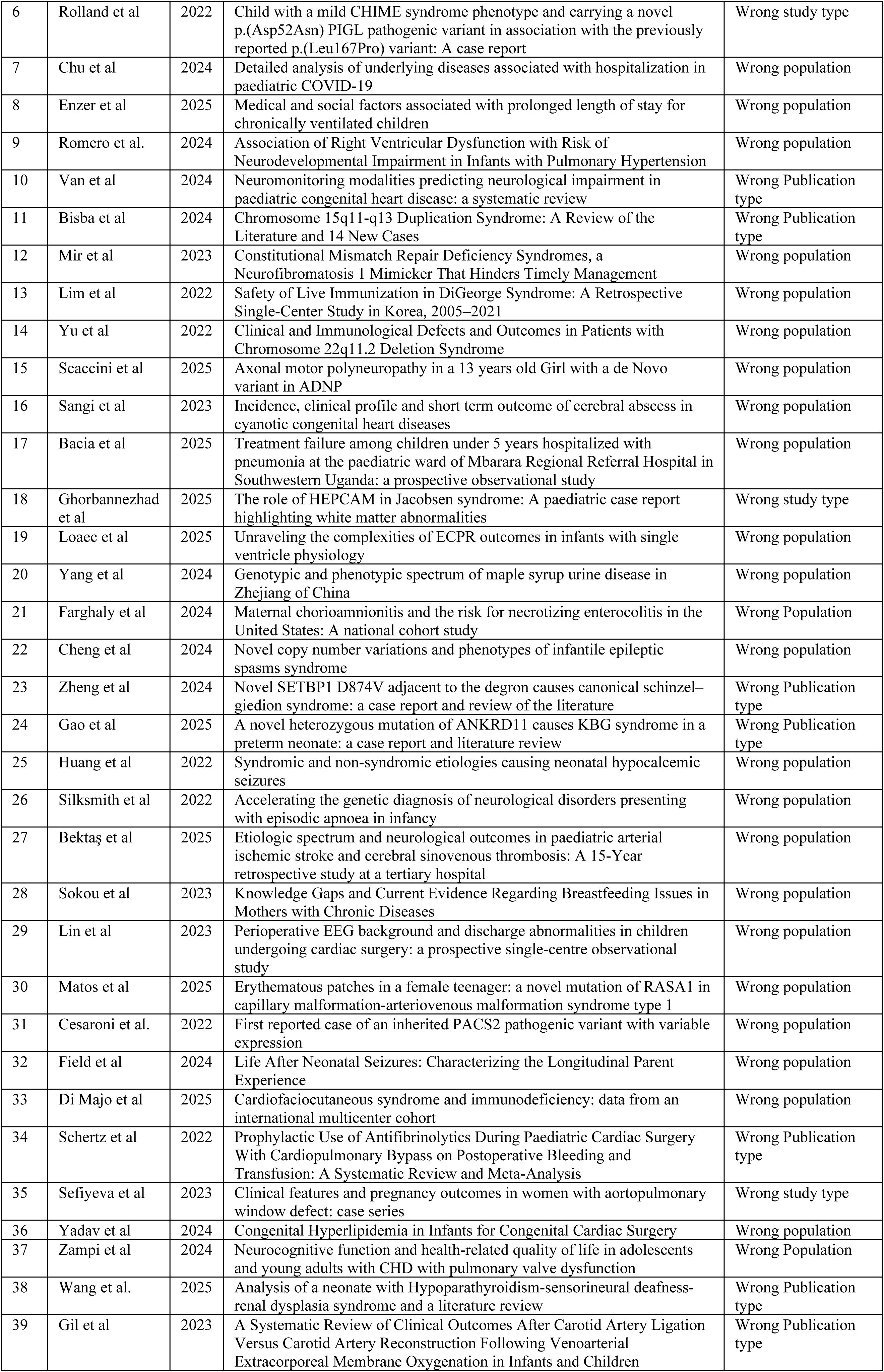
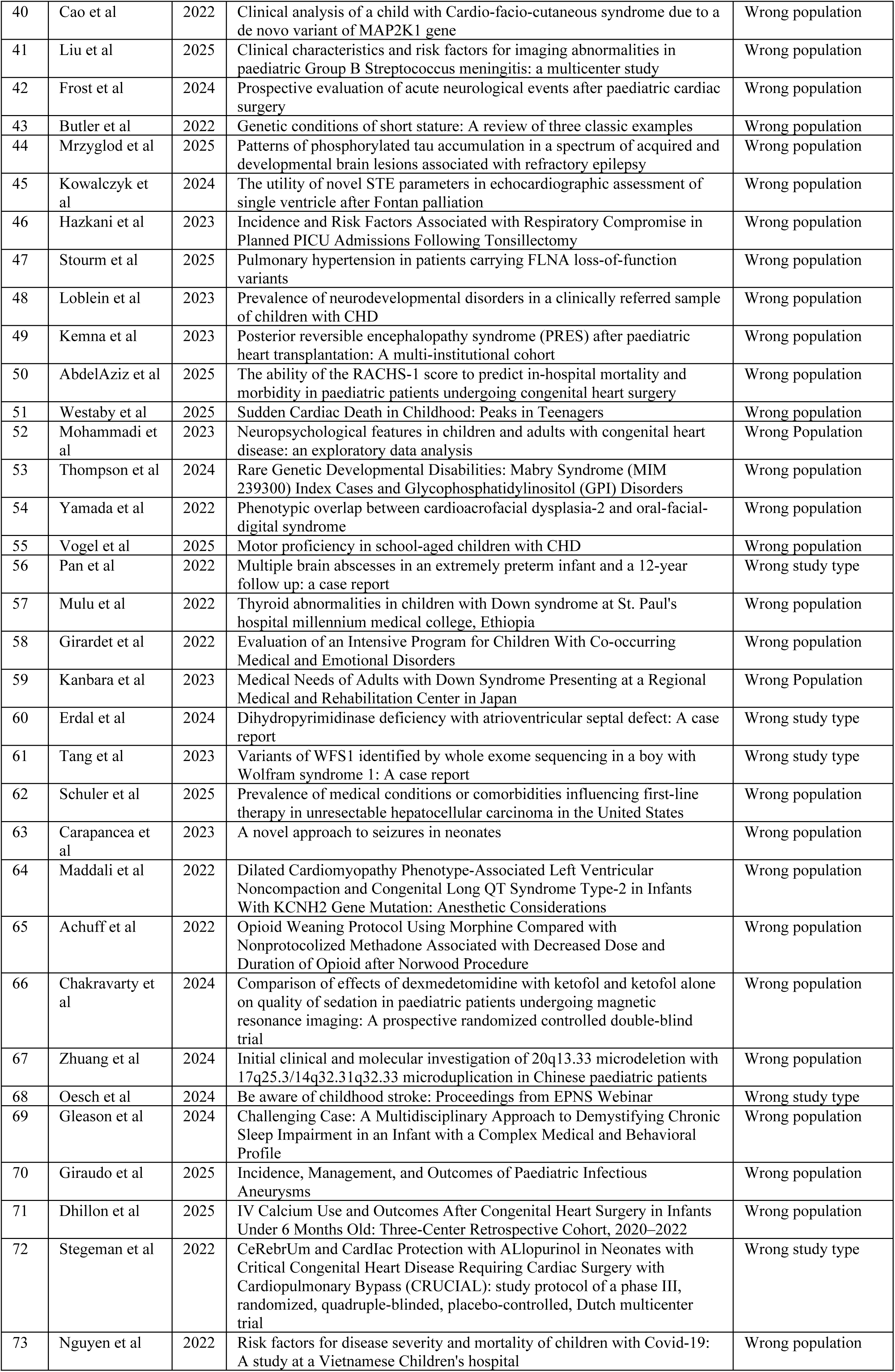
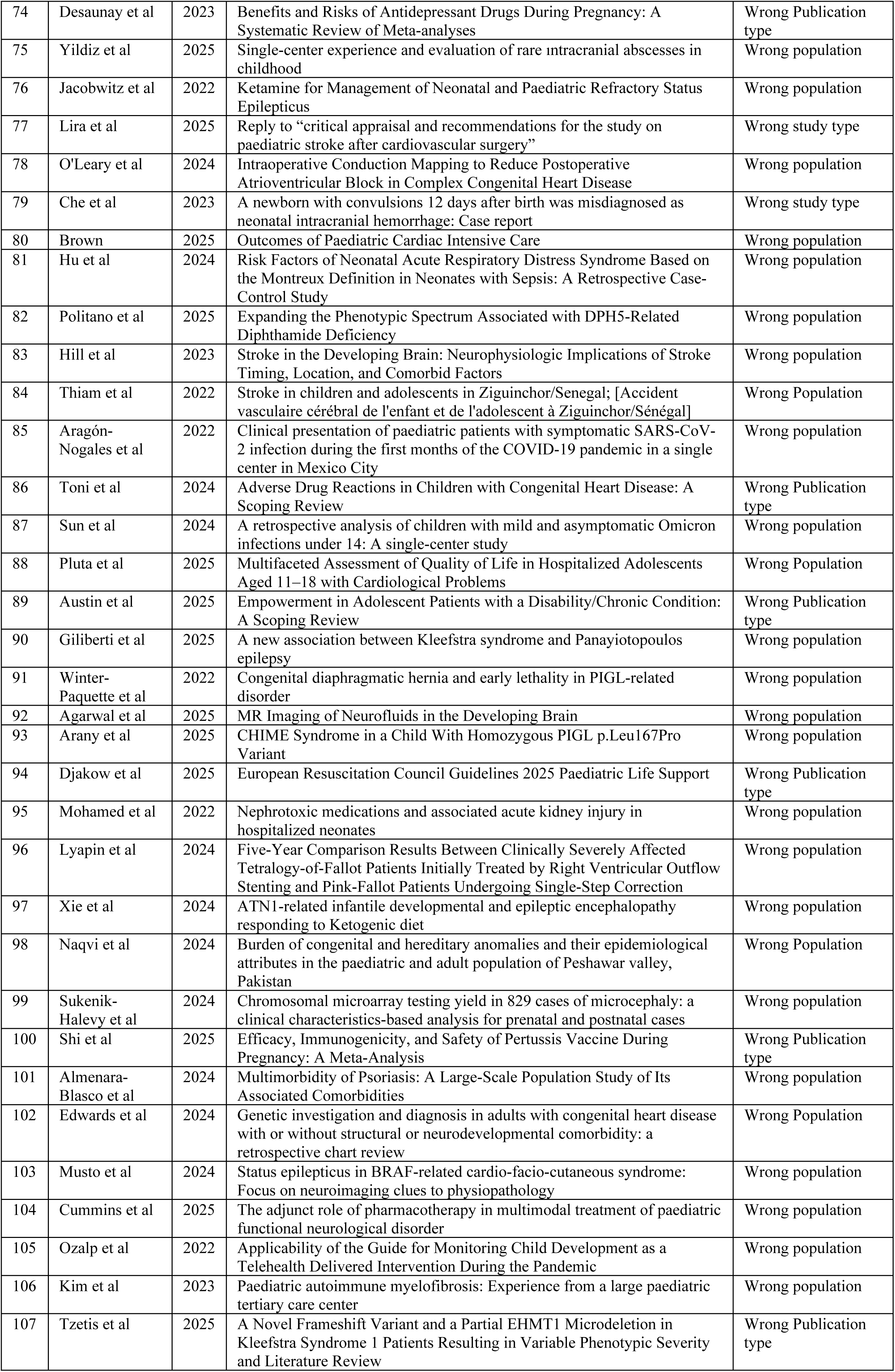
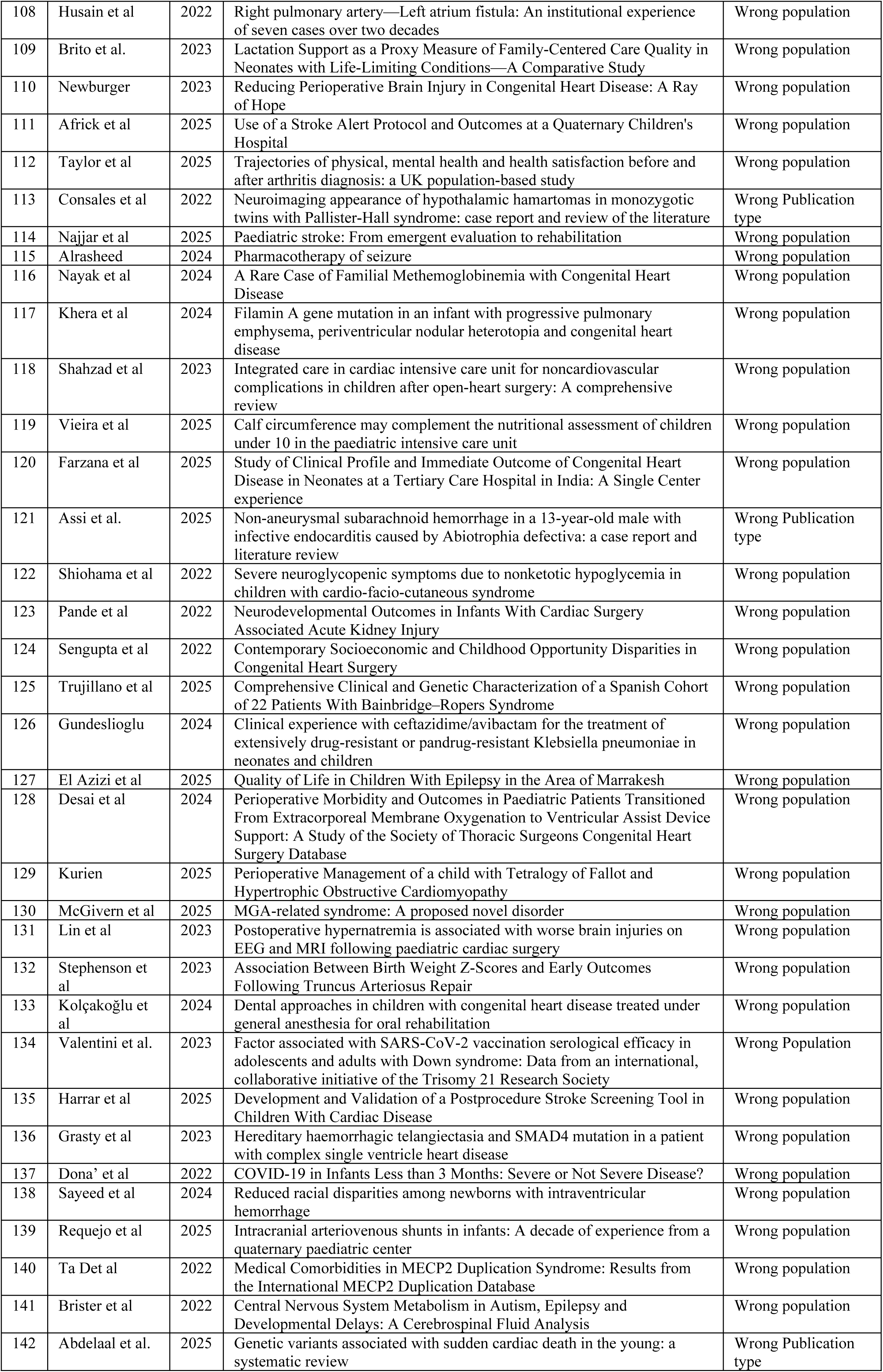
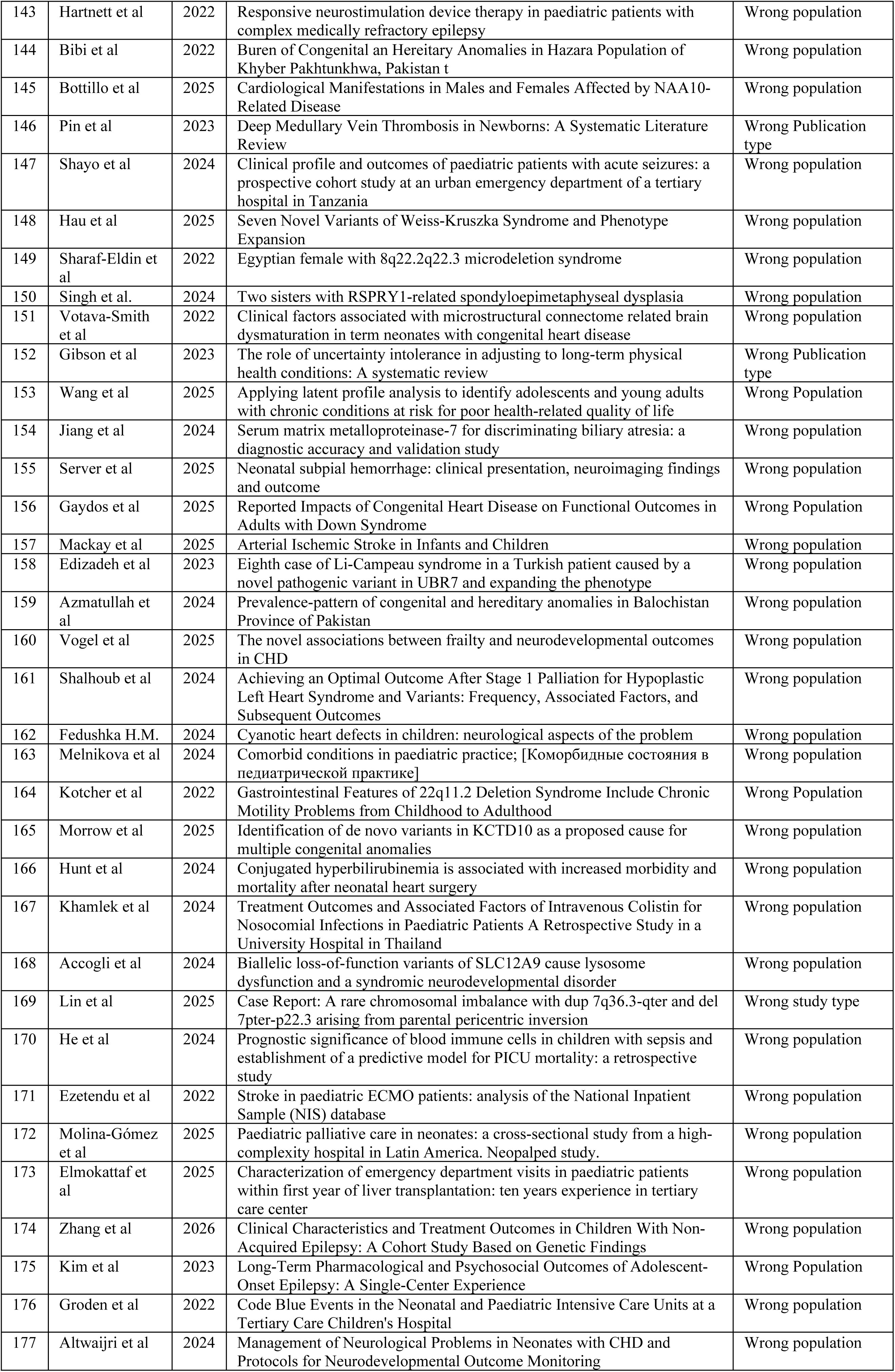
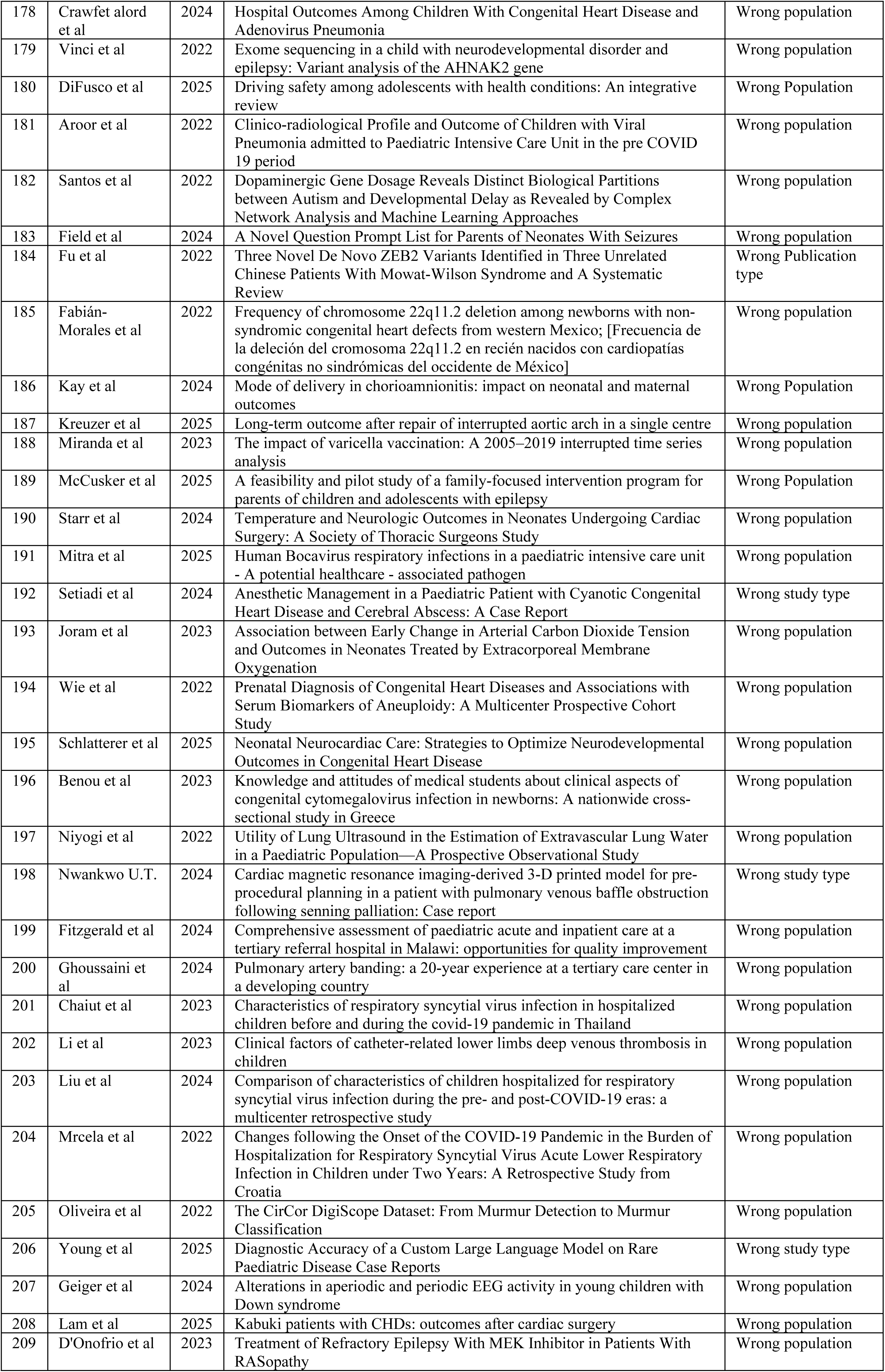
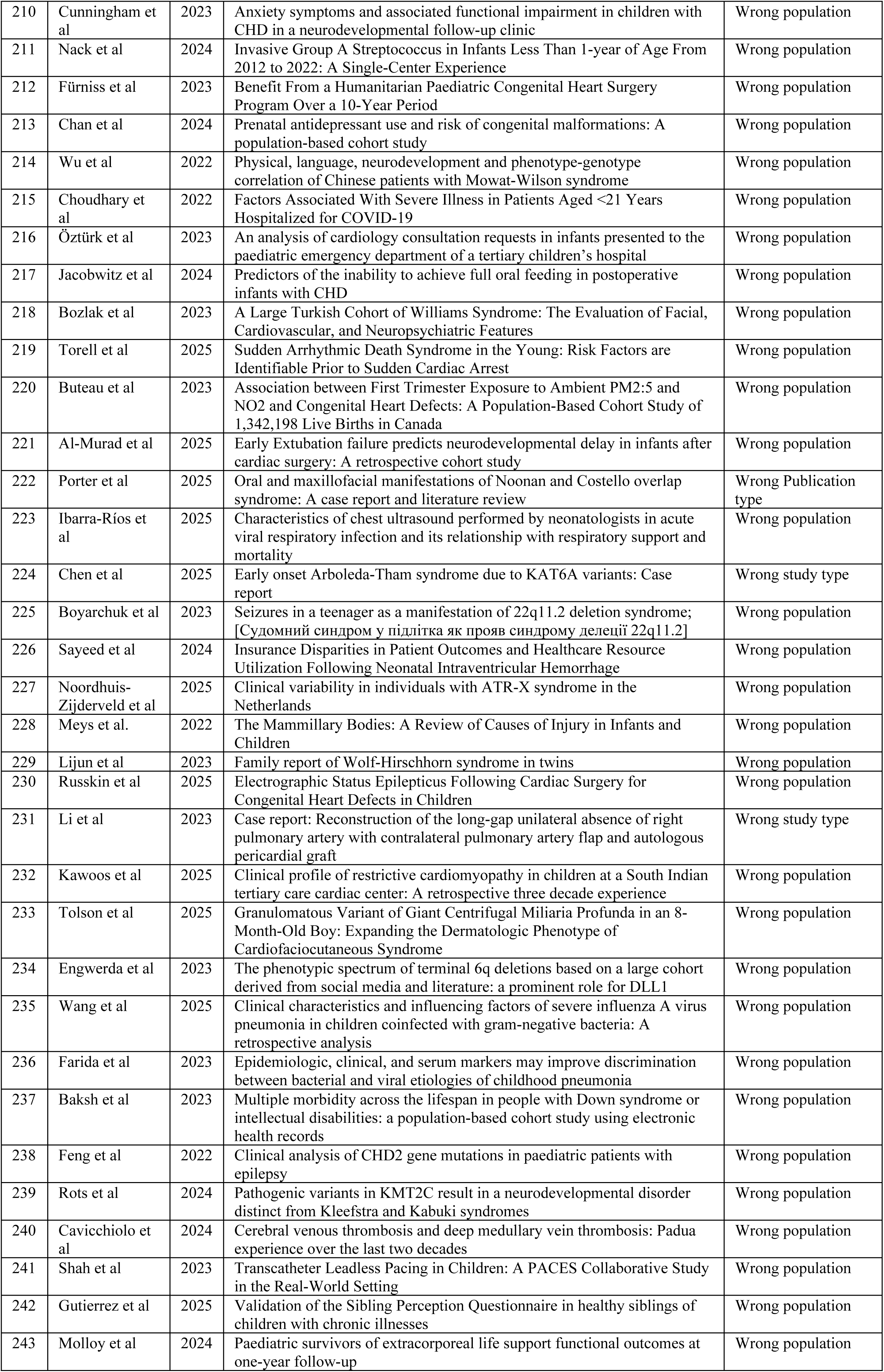
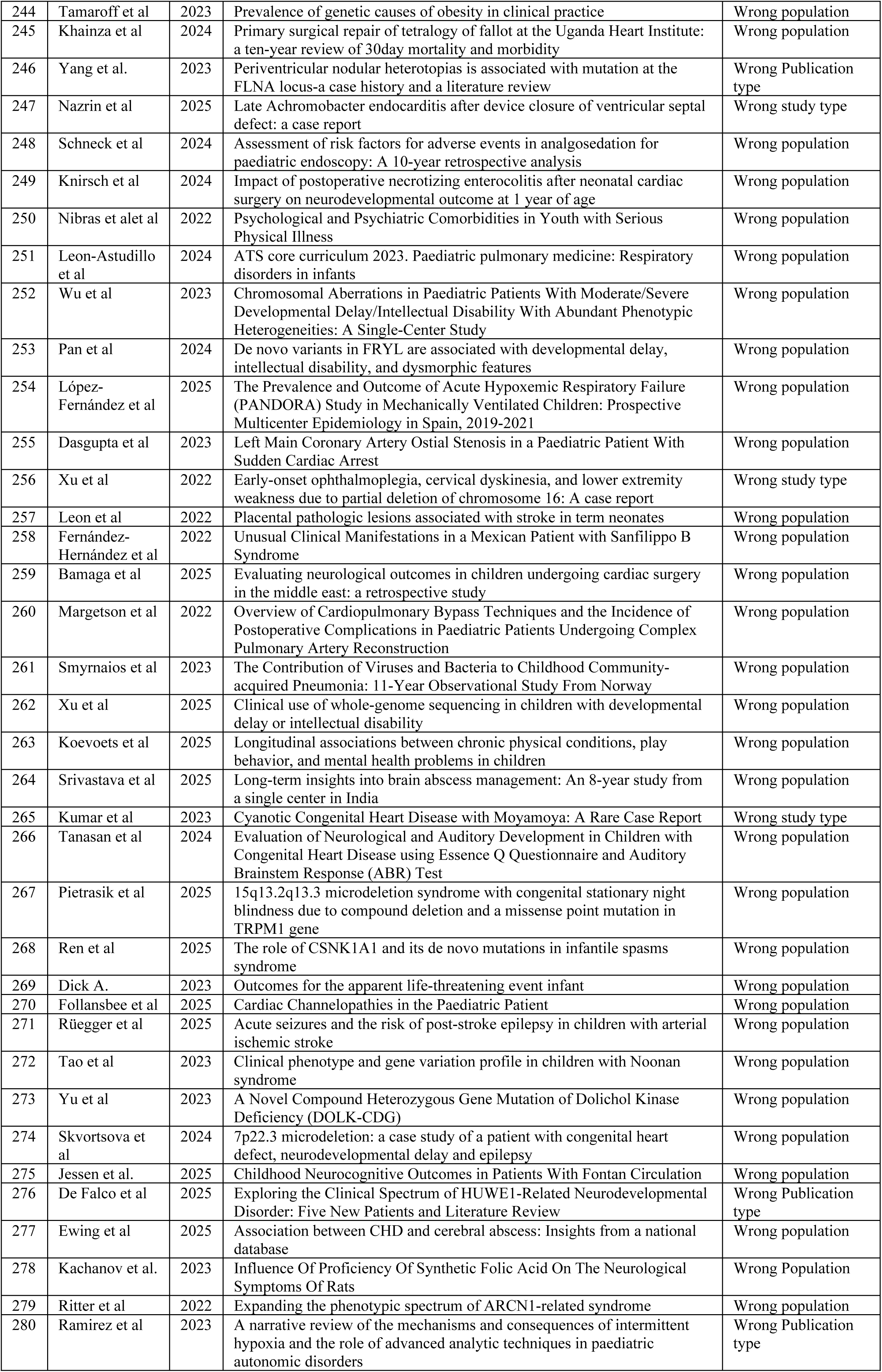
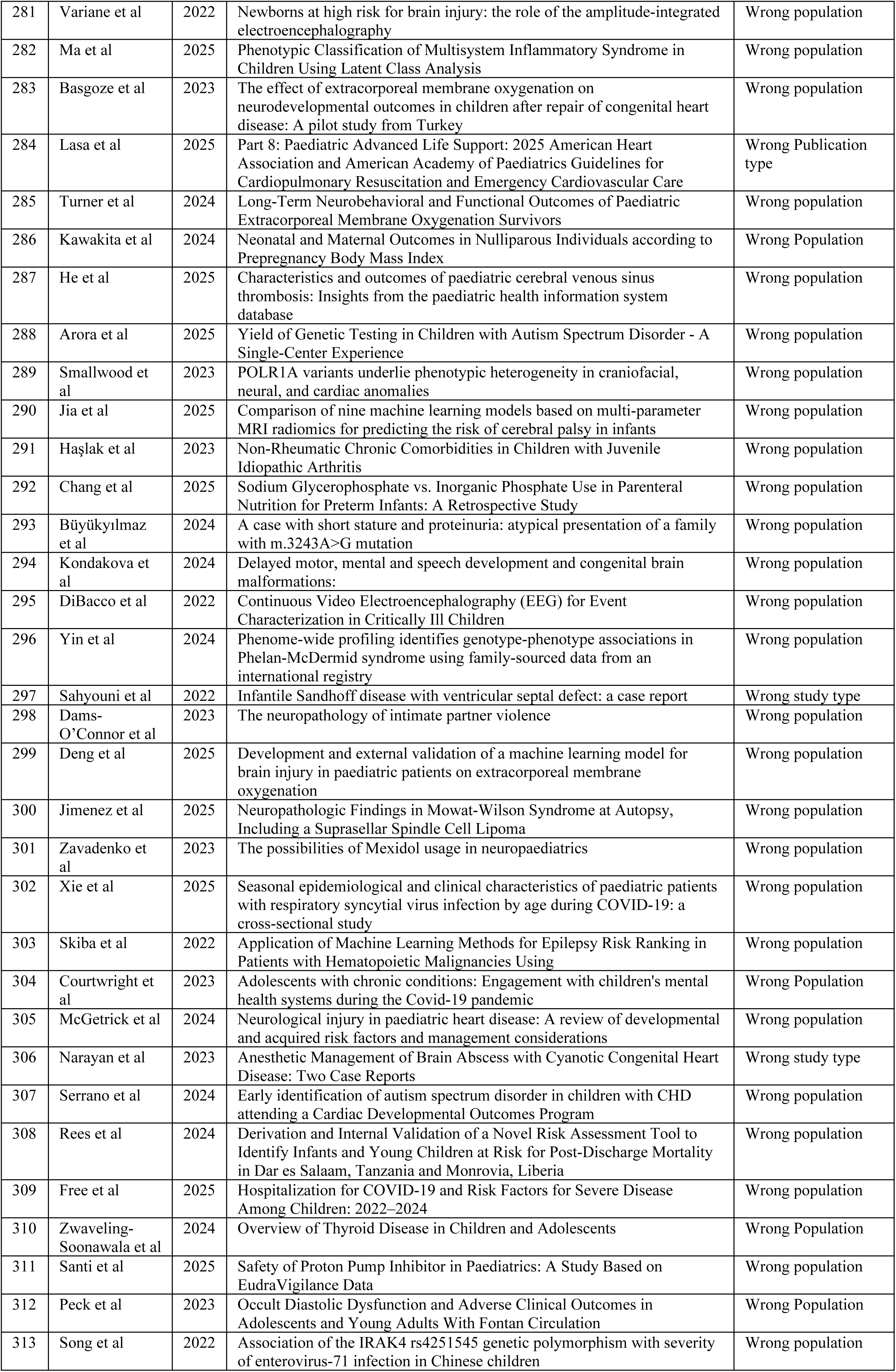
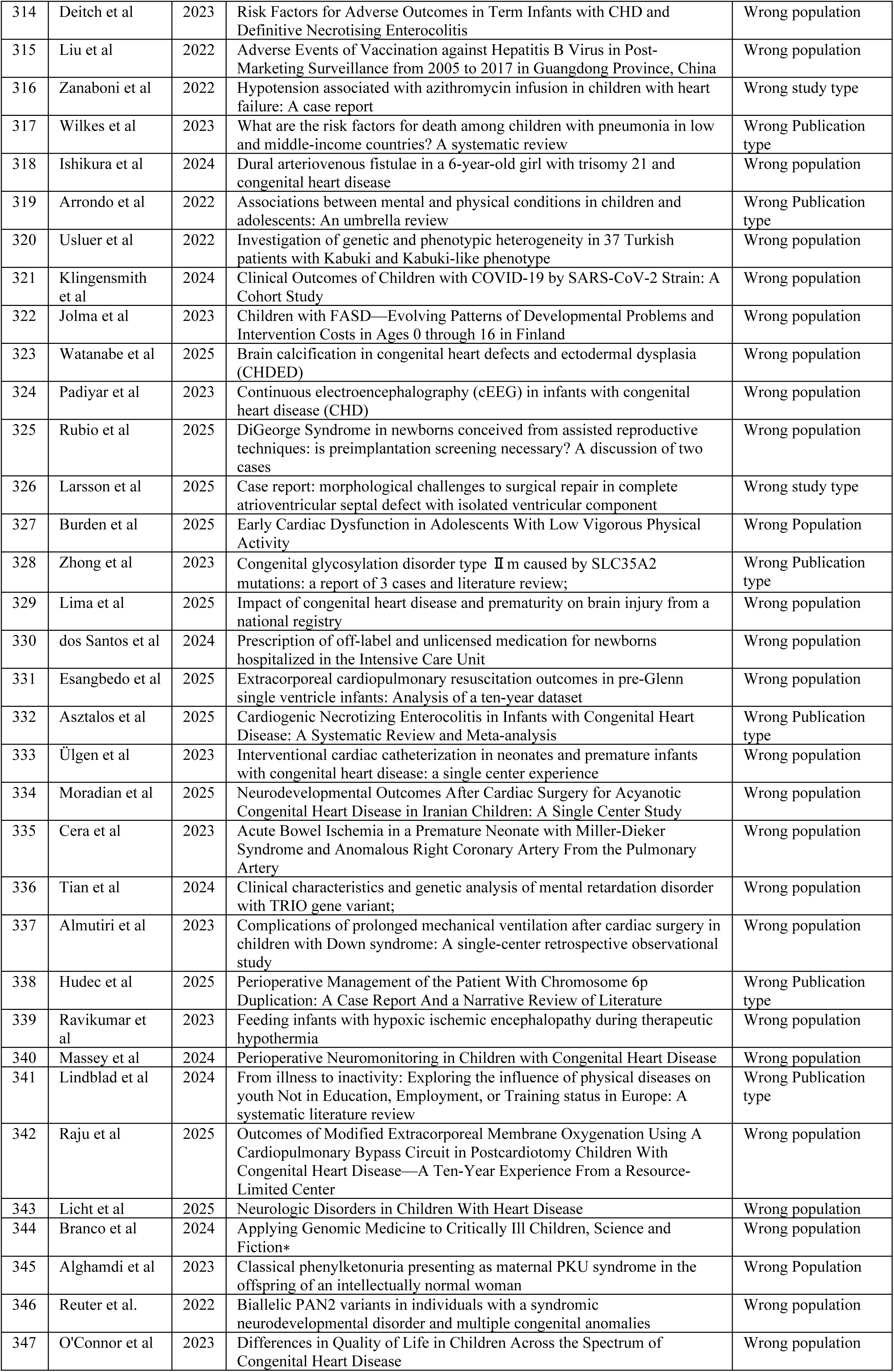
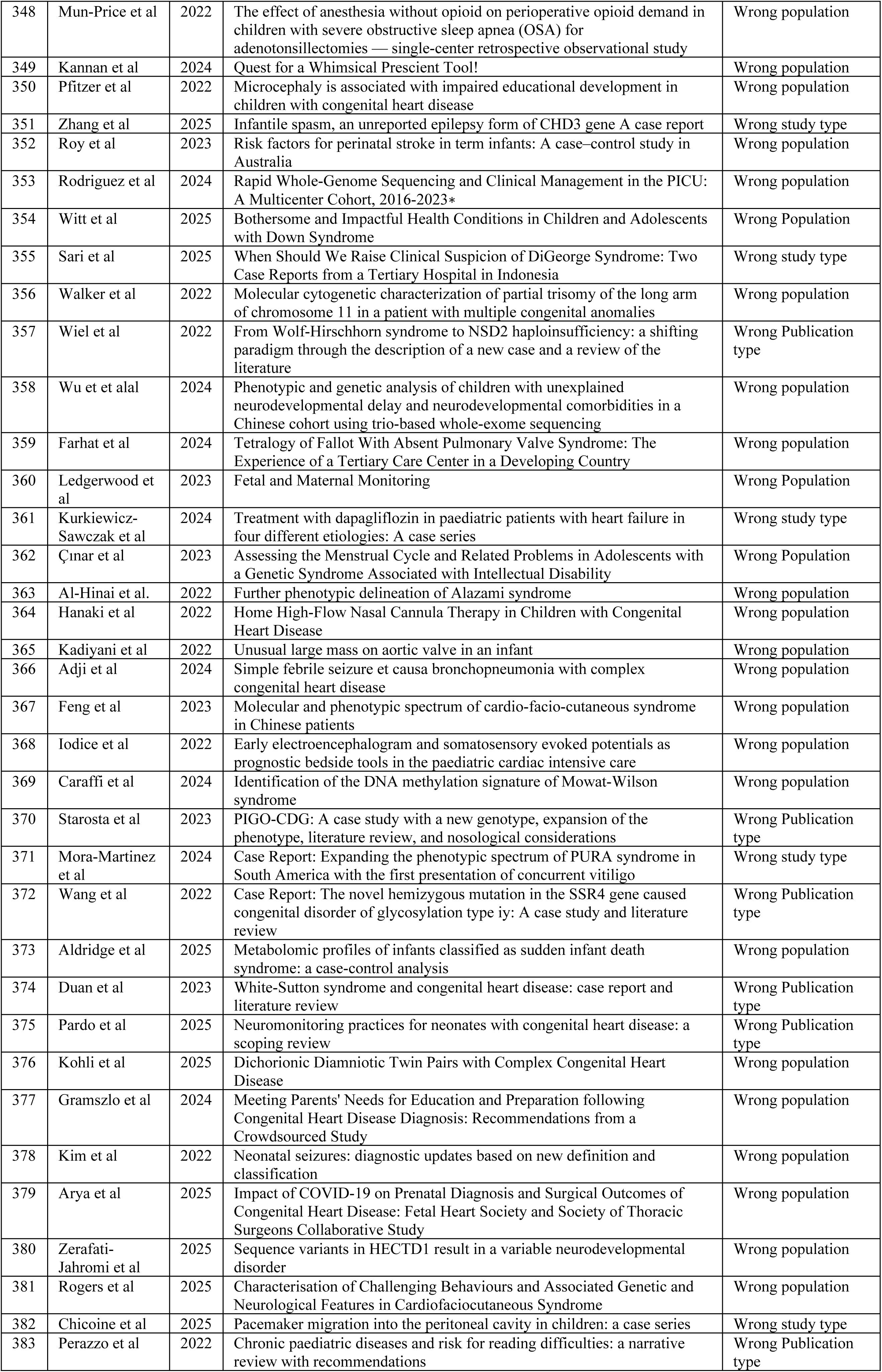
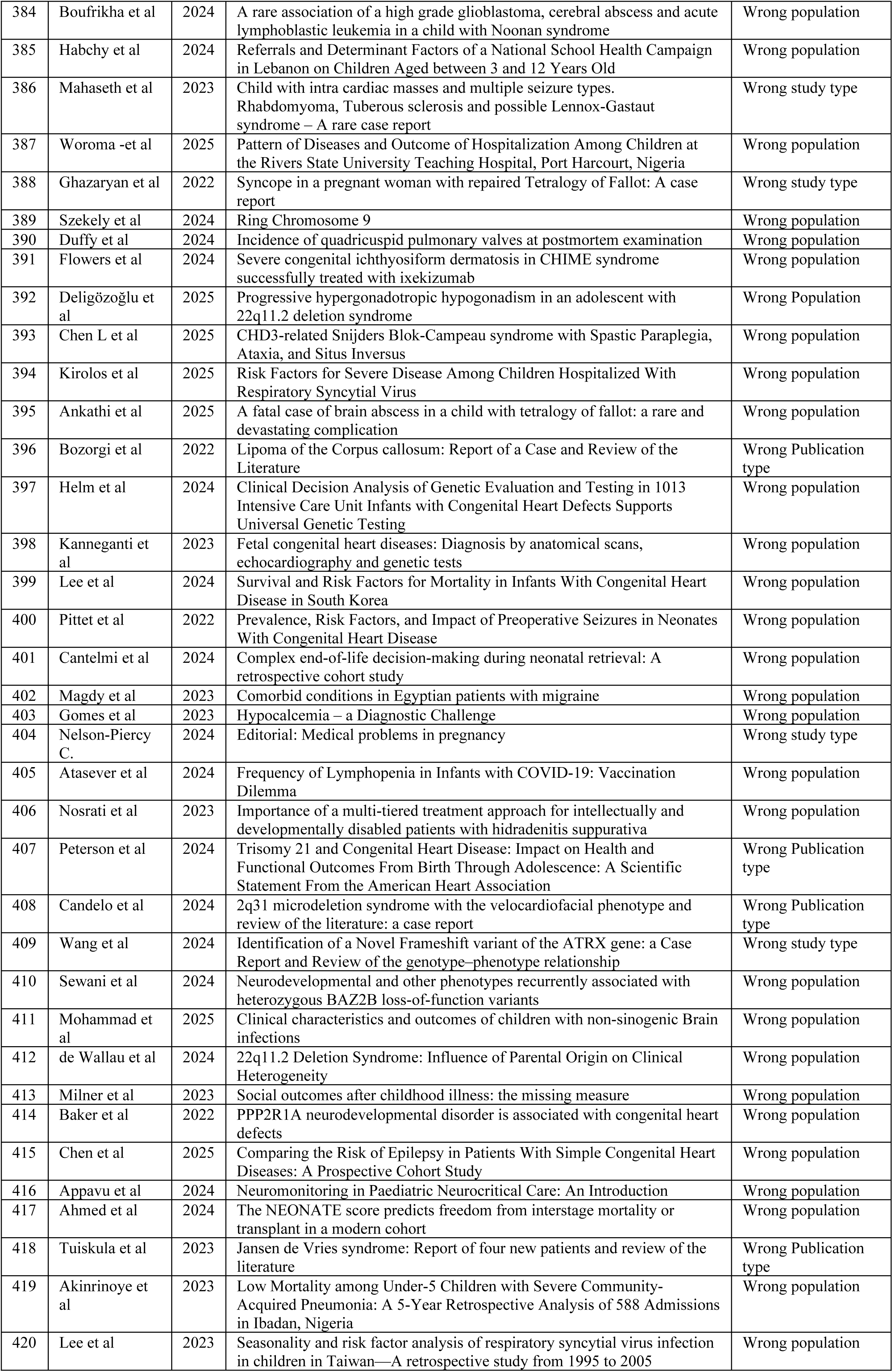
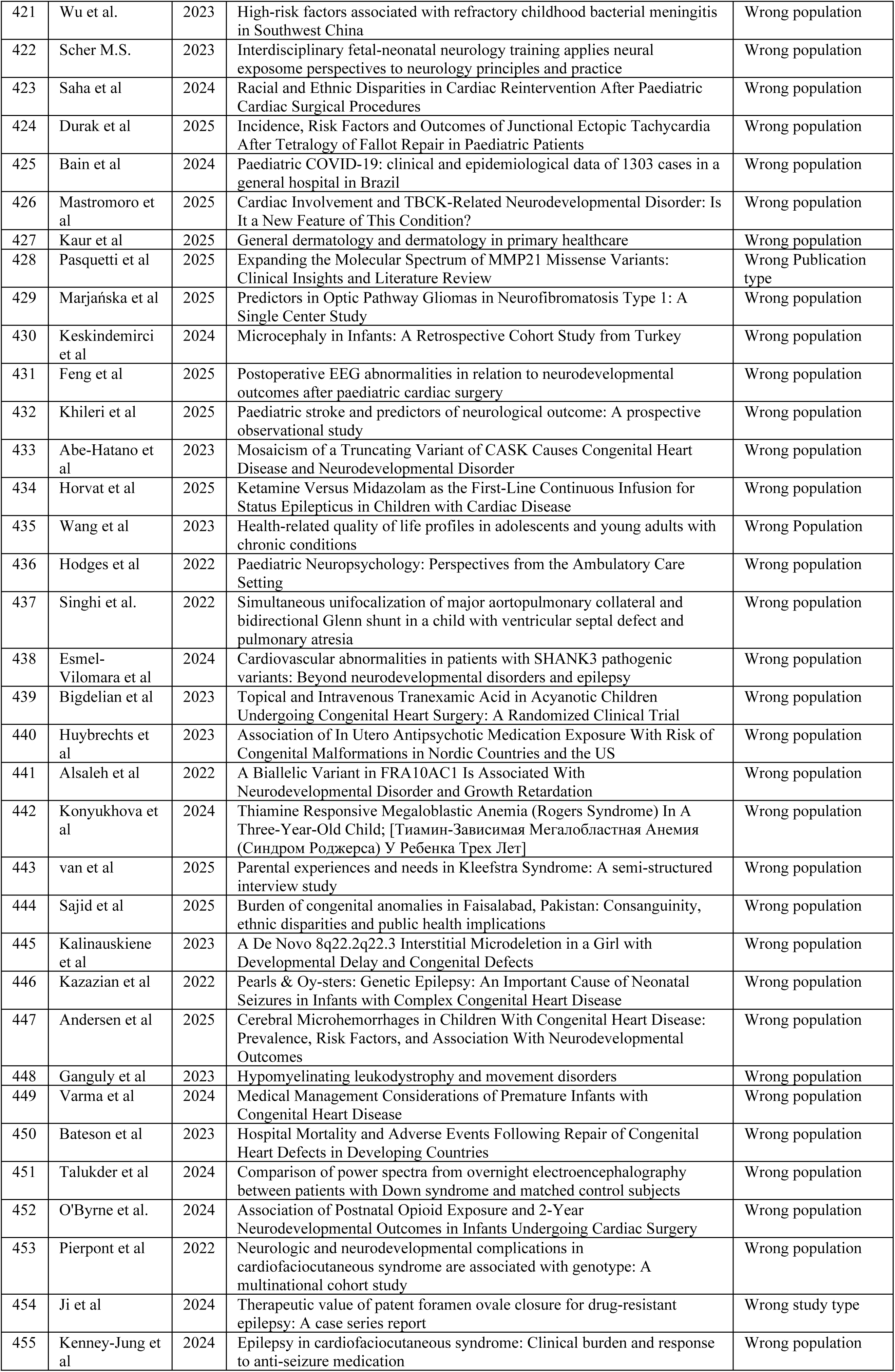
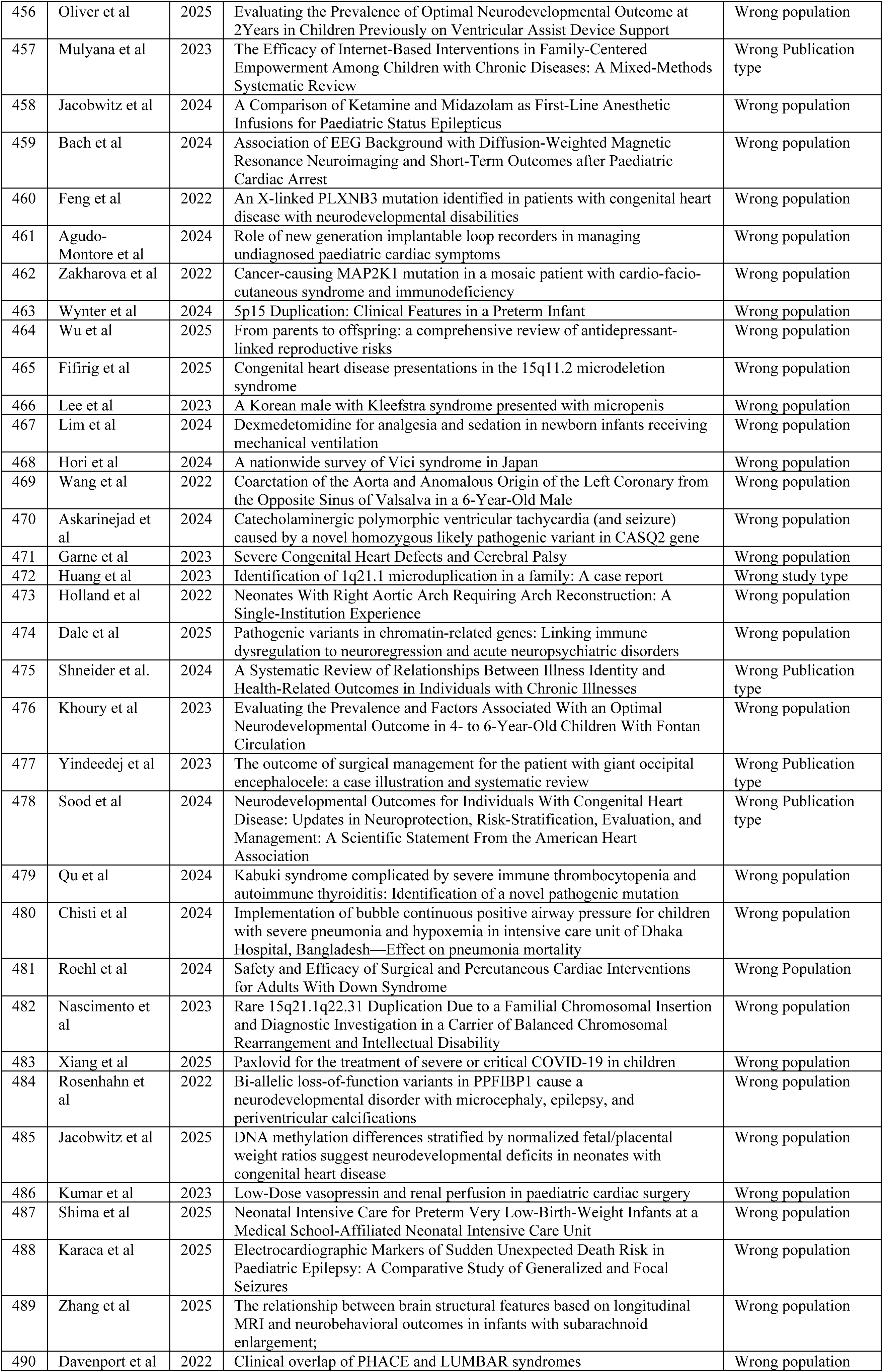
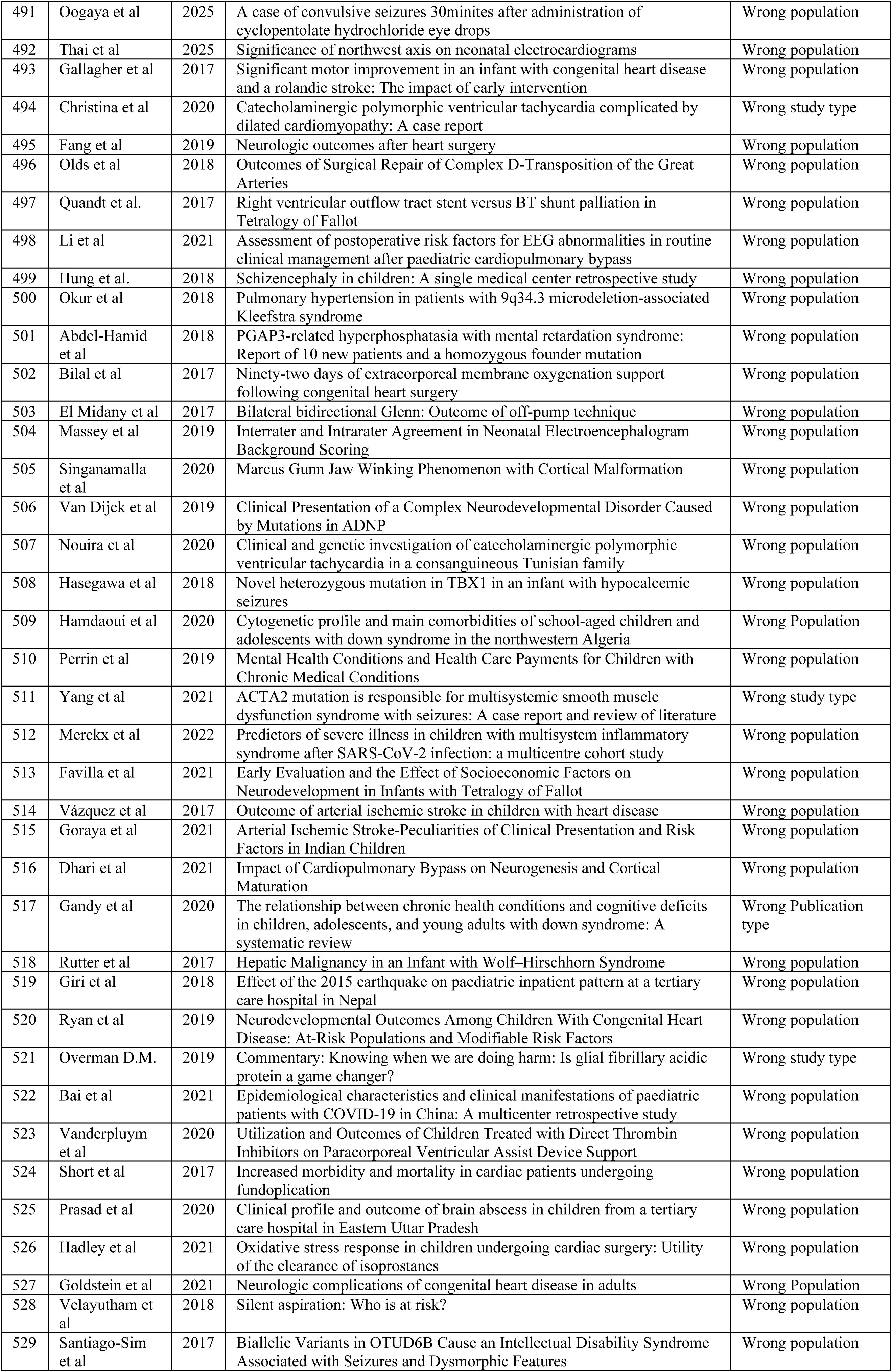
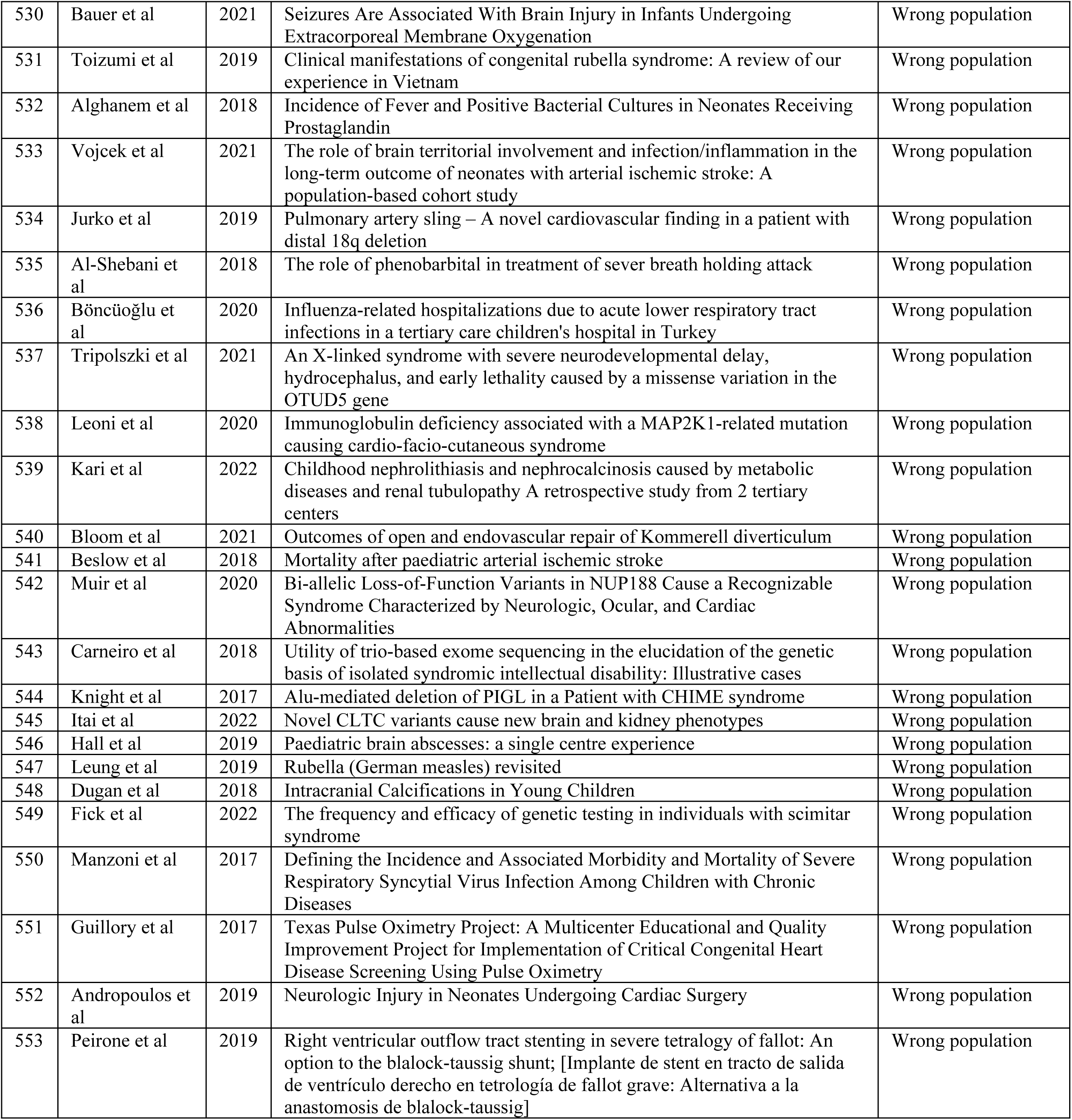
Excluded Studies and Reasons for Exclusion.

**Supplementary File 3:** Inter-rater Reliability Assessment.

| References | Accessors | Q1 | Q2 | Q3 | Q4 | Q5 | Q6 | Q7 | Q8 | Q9 |
| --- | --- | --- | --- | --- | --- | --- | --- | --- | --- | --- |
| Massin et al. [21] | Reviewer A | 1 | 1 | 1 | 1 | 1 | 1 | 1 | 1 | 1 |
|  | Reviewer B |  |  |  |  |  |  |  |  |  |
| Desnoux et al. [20] | Reviewer A | 1 | 1 | 0 | 1 | 1 | 1 | 1 | 1 | 0 |
|  | Reviewer B |  |  |  |  |  |  |  |  |  |
| Castillo-Pinto et al. [5] | Reviewer A | 1 | 1 | 0 | 1 | 0 | 1 | 1 | 1 | 0 |
|  | Reviewer B |  |  |  |  |  |  |  |  |  |
| Chen et al. [23] | Reviewer A | 1 | 1 | 1 | 1 | 1 | 1 | 1 | 1 | 0 |
|  | Reviewer B |  |  |  |  |  |  |  |  |  |
| Leisner et al. [22] | Reviewer A | 1 | 1 | 1 | 1 | 1 | 1 | 1 | 1 | 1 |
|  | Reviewer B |  |  |  |  |  |  |  |  |  |
| Ghosh et al. [4] | Reviewer A | 1 | 1 | 0 | 1 | 0 | 1 | 0 | 1 | 0 |
|  | Reviewer B |  |  |  |  |  |  |  |  |  |
| Shibata et al. [6] | Reviewer A | 1 | 1 | 1 | 1 | 1 | 1 | 1 | 1 | 0 |
|  | Reviewer B |  |  |  |  |  |  |  |  |  |
| Ikegawa et al. [24] | Reviewer A | 1 | 1 | 1 | 1 | 1 | 1 | 1 | 1 | 0 |
|  | Reviewer B |  |  |  |  |  |  |  |  |  |
| Percentage Agreement | Overall |  |  |  |  |  |  |  |  |  |

Q1: Was the sample frame appropriate to address the target population?
Q2: Were study participants sampled appropriately?
Q3: Was the sample size adequate?
Q4: Were study subjects and settings described in detail?
Q5: Was the data analysis conducted with sufficient coverage?
Q6: Were valid methods used for identification of epilepsy?
Q7: Was epilepsy measured in a reliable and standardised manner?
Q8: Was appropriate statistical analysis performed?
Q9: Was the response rate adequate and appropriately managed?

## Notes

### Competing Interest Statement

The authors have declared no competing interest.

### Funding Statement

The author(s) received no specific funding for this work.

